# Leapfrogging cancer diagnostics with AI: a mixed-methods assessment of implementation determinants in low- and middle-income countries

**DOI:** 10.64898/2026.10.19.26365881

**Authors:** Lydia A. Schoenpflug, Nikki J. van den Berg, Serey Vathana Chhut, Elaine Y. L. Leung, Lai-Meng Looi, Robert Lukande, Samuel James Meja, Rafael Parra-Medina, Tilak Pathak, Jeremia J. Pyuza, Rubina Razack, Augustine Tawiah, Maureen W. Waithaka, Pablo Zoroquiain, Jurriaan Barkey Wolf, Marcus J. Rijken, Gernot Jundt, Maxime W. Lafarge, Tjalling Bosse, Viktor H. Koelzer, Nanda Horeweg

## Abstract

**Introduction:** Cancer incidence is rising rapidly in low- and middle-income countries (LMICs), where diagnostic pathology capacity is constrained. AI applied to routine histopathology could strengthen diagnostic capacity and offer leapfrogging potential by embedding AI into diagnostic systems as they are built up. However, without empirically identifying implementation determinants in LMIC cancer-diagnostic settings, AI solutions risk being poorly aligned with local constraints, limiting impact or exacerbating existing disparities.

**Methods:** We conducted an exploratory mixed-methods pilot study across three phases: 1) a structured survey characterising the diagnostic and technical landscape and AI priorities; 2) focus group discussions (FGDs) to identify implementation determinants and 3) a second survey quantifying FGD-derived themes. Twelve physicians (nine pathologists, three gynaecologists) were purposively recruited (10 LMIC, 2 high-income countries for reference) and findings integrated through narrative triangulation.

**Results:** Diagnostic and AI infrastructure varied substantially and did not track consistently with country income classification. H&E staining was universally available, immunohistochemistry capacity was below WHO-recommended capacity for cancer diagnosis at 7 of 10 sites and molecular testing absent or minimal at 6 of 10 sites. Whole-slide-imaging scanners were available at 8 of 10 sites, graphics processing units, required for efficient AI inference, were available at only 4 of 10 sites. Participants prioritised AI for cancer screening, diagnosis, and biomarker prediction from H&E slides. Six implementation determinants were identified: AI literacy, governance frameworks, funding, evidence generation, clinical deployment, and collaboration. Financial constraints emerged as the dominant barrier across all determinants, and collaboration as a key enabler.

**Conclusion:** AI implementation readiness varied substantially within income groups, suggesting that country income classification is an insufficient proxy for institutional readiness, underscoring the need for institution-level assessment. Realising the leapfrogging potential of AI in this context requires coordinated progress across all determinants, with sustainable financing, and collaboration as enablers of the other determinants.

**Key messages:** *What is already known on this topic?:* Cancer incidence is rising rapidly in LMICs, where diagnostic capacity and molecular testing capabilities are severely constrained. AI could alleviate gaps, yet evidence on implementation determinants for integrating AI into cancer diagnostics in these settings remains limited.

*What this study adds?:* In this exploratory sample, AI implementation readiness varied substantially across LMIC settings, including within the same country income group. Readiness is shaped by six implementation determinants: AI literacy, governance frameworks, funding, evidence generation, clinical deployment, and collaboration, with financial constraints as the dominant cross-cutting barrier.

*How this study might affect research, practice or policy?:* These findings support institution-level feasibility assessment rather than reliance on country income classification as a proxy and highlight the need for sustainable financing and collaboration as foundational prerequisites for AI implementation in cancer diagnostics in LMICs.

## Introduction

Cancer incidence and mortality are rising rapidly in low- and middle-income countries (LMICs), following trends previously observed in high-income countries (HICs) but with disproportionately poorer outcomes (1). Effective cancer care depends on timely detection and accurate diagnosis, which guides prognostic assessment and informs treatment selection. Many LMICs face critical shortages in cancer diagnostic capacity, including limited diagnostic infrastructure, lack of pathologists, and geographical concentration of expertise in tertiary public hospitals (2). Manual workflows and inconsistent quality assurance further compound these constraints. As a result, patients frequently experience postponed treatment initiation and suboptimal care for time-sensitive or high-burden malignancies, reinforcing inequities in cancer outcomes (3,4).

Despite major advances in digital pathology, molecular diagnostics, and targeted therapies, access to these innovations remains limited in LMIC settings. Artificial intelligence (AI) has emerged as a potential strategy to alleviate this gap: AI models have demonstrated the ability to perform cancer screening, detection, diagnosis, and prediction of molecular biomarkers directly from hematoxylin and eosin (H&E)-stained slides (5,6), one of the most widely available diagnostic resources across LMIC health systems. Further, growing interest in AI capabilities and their health applications has prompted policy responses at the global and regional level, with the WHO issuing guidance on the ethics and governance of AI for health (10) and the African Union adopting a Continental Artificial Intelligence Strategy in 2024 in which health is named as a priority sector. Taken together, these developments provide momentum for leapfrogging: rather than replicating the incremental development path of HIC settings, LMICs could design cancer diagnostic systems with AI embedded from the outset, using it to triage cases, optimise allocation of resource-intensive tests such as immunohistochemistry (IHC) or molecular diagnostics, and improve the efficiency of existing and developing clinical capacity (7–9). However, whether AI can realistically contribute to more equitable cancer diagnostics in LMICs remains uncertain: available algorithms cover a limited portfolio of applications, and their applicability depends on the implementation readiness of individual settings. Without systematic identification and assessment of the determinants shaping this readiness, AI solutions risk being poorly aligned with local health system constraints, potentially limiting impact or exacerbating the global disparities they aim to address.

Existing evidence on AI implementation in cancer diagnostics predominantly reflects HIC experiences (11–13) or broader qualitative evidence bases in which LMIC settings remain substantially underrepresented (14). Among studies engaging with LMIC contexts, most are limited to expert viewpoints from a narrow geographic base (15,16), clinical infrastructure assessment (17), or broad literature syntheses (18,19). Critically, none have empirically identified or evaluated the implementation determinants shaping AI adoption in practice, including governance readiness, clinician and public attitudes towards AI, funding structures, stakeholder roles, and collaboration, leaving the determinants of AI implementation readiness in LMIC cancer-diagnostic settings largely uncharacterised.

This mixed-methods pilot study addresses this gap by (1) characterizing the diagnostic and AI infrastructure landscape, (2) identifying the determinants relevant to AI implementation for cancer diagnostics and (3) quantifying these determinants, using focus group discussions and surveys with participants from ten LMIC settings.

## Methods

### Study Design

This mixed-methods pilot study was conducted in three phases and reported in accordance with the Good Reporting of a Mixed Methods Study (GRAMMS) framework. Quantitative components are additionally reported in line with the Checklist for Reporting Results of Internet E-Surveys (CHERRIES), and qualitative components in accordance with the Consolidated Criteria for Reporting Qualitative Research (COREQ) (**Supplementary Files 1-3**).

The study comprised three sequential phases: Survey 1 (October-November 2025), focus group discussions (FGDs; January-February 2026), and Survey 2 (June 2026), reflecting an exploratory sequential mixed-methods design (20). Survey 1 characterised the current diagnostic landscape, including digitisation capacity, laboratory infrastructure, and workforce availability, and captured participants’ expressed needs and priorities for AI, using a structured quantitative instrument. FGDs were subsequently conducted to identify the determinants relevant to AI implementation in this study population. Themes identified in the FGDs were then operationalised into Survey 2, allowing systematic quantification of the determinants identified qualitatively. A small number of HIC participants were included in the study for contextual comparison. Full Survey 1 and Survey 2 instruments are provided in **Supplementary Files 4 and 7**, all Survey responses are provided in **Supplementary Files 6 and 8**.

### Ethical considerations

The study was exempted from formal ethics review by the Ethics Committee Northwest and Central Switzerland (EKNZ) (Req-2026-00111). All participants provided informed electronic consent.

### Patient and public involvement

Patients and the public were not involved in the design, conduct, reporting, or dissemination of this study.

### Study setting and recruitment

Participants were purposively recruited from professional networks and through targeted identification of physicians (9 pathologists, 2 gynaecologists, 1 gynaecological oncologist) involved in cancer diagnostics and digital pathology research. Selection aimed to include one respondent per country and to reflect a range of country income levels and geographic regions, spanning Africa, Asia, Europe and South America. Of 15 individuals approached, 13 participated in at least one study component. One participant was excluded due to a protocol deviation, resulting in a final analytic sample of 12 participants (10 LMIC, 2 HIC included as illustrative reference points) across 12 countries. All participants were subsequently invited to co-author this manuscript. Data collection and primary analysis were completed by non-participant investigators.

### Study procedures

#### Survey 1

Survey 1 comprised seven domains: participant characteristics, laboratory infrastructure, pathology workforce, oncology treatment availability, AI applications, AI requirements and feasibility of AI implementation. Items were developed iteratively and reviewed by a regional expert with LMIC experience (MG). Participants completed the survey via Google Forms, with one of two researchers (LAS, NJvdB) present via Zoom guiding participants through the form and clarifying items with a standardised script (**Supplementary File 5**).

#### Focus group discussions

FGDs informed interpretation of Survey 1 findings and explored implementation readiness and pathways, using a semi-structured guide (**Supplementary File 9**) covering implementation steps, stakeholder roles, governance, and collaboration, with additional probes on regulatory, and ethical barriers. Nine participants took part across three sessions (n=6, 2, 1). Sessions (60-90 minutes) were conducted online in English via secure video-conferencing, audio-recorded with consent, and facilitated by NH (clinical epidemiologist, PhD). One to three non-participating study team members attended as observers, no field notes were taken. Two participants had prior academic collaboration with the study team, all others were unknown to the investigators prior to recruitment. Recordings were transcribed using AI-assisted transcription (Whisper v3 (21)) and manually verified by LAS, transcripts were not returned to participants for review.

#### Survey 2

Survey 2, provided to all participants from the prior two phases, assessed perceived implementation readiness across five domains: AI awareness and attitudes, AI literacy and education, AI governance, ethics and funding, stakeholders and collaboration, and clinical deployment, derived from FGD findings and refined by investigator consensus (LAS, NH). The survey was self-administered electronically via Google Forms without interviewer involvement (approx 15 minutes).

### Data Analysis

Survey data were analysed descriptively using Excel and Python. Two duplicate Survey 2 submissions were identified; in both cases, the most recent response was retained for analysis.

To enable visual comparison of infrastructure levels across participants, descriptive composite scores were developed by assigning points to Survey 1 infrastructure items. Clinical infrastructure scoring included IHC capacity (1: limited daily; 2: limited weekly; 3: sufficient), molecular testing availability (0: none; 1: PCR/HPV DNA; +1 for additional modalities, maximum 5), and pathology workforce capacity (1: few general pathologists; 2: general pathologists; 3: general and subspecialty-trained pathologists). AI infrastructure scoring assigned points to: whole-slide image (WSI) scanners (0: none; 1: low-throughput; 2: high-throughput), and 1 point for presence of each of the following: microscope cameras, CPU servers, graphical processing units (GPUs), GPU servers and cloud storage.

Focus group transcripts were coded in QualCoder v3.7 (22) using a hybrid inductive-deductive thematic analysis approach informed by the TICD framework for determinants of healthcare professional practice (23). LAS developed an initial code tree, which was refined jointly with NH, the final code book can be found in **Supplementary File 10**. LAS and NH independently coded all transcripts. Raw percent agreement across codes was high (median 98.6%, range 93.8-99.9%), while Cohen’s kappa values were lower and more variable (median 0.18, range -0.04 to 0.69), consistent with known sensitivity of kappa to low-prevalence codes in thematic datasets. Discrepancies were resolved through discussion, with iterative refinement of the codebook and a final reconciled coding dataset used for analysis. The research team’s interest in AI implementation in LMICs may have introduced implicit framing toward the topic’s potential, this was mitigated through open-ended, non-leading questions. All participants reviewed the manuscript as co-authors, providing an additional form of respondent validation.

Integration of quantitative and qualitative findings occurred through narrative comparison during interpretation. Survey 1 findings informed the development of the focus group guide, and FGD-derived themes informed the construction of Survey 2, enabling iterative triangulation across study phases.

## Results

Results are presented across two surveys and FGDs. Quantitative results refer to LMIC participants (n=10) unless otherwise stated; results from HIC participants (n = 2) are noted explicitly.

### Participant characteristics

Characteristics of the 12 participants are presented in **Table 1**. All 12 participants completed Survey 1 and Survey 2, and 9 (75%) took part in the FGDs.

**Table 1:** Participant characteristics.

| **Participant characteristic** | **n** | **%** |
|---|---|---|
| **Country income level** |  |  |
| LMIC | 10 | 83 |
| - Low-income (Malawi, Uganda) | 2 | 17 |
| - Lower-middle-income (Cambodia, Ghana, Kenya, Nepal, Tanzania) | 5 | 42 |
| - Upper-middle-income (Colombia, Malaysia, South Africa) | 3 | 25 |
| HIC* (Chile, UK) | 2 | 17 |
| **Region** |  |  |
| Africa | 6 | 50 |
| Asia | 3 | 25 |
| South America | 2 | 17 |
| Europe | 1 | 8 |
| **Gender** |  |  |
| Male | 8 | 67 |
| Female | 4 | 33 |
| **Profession** |  |  |
| Pathologist | 9 | 75 |
| Gynaecologist | 2 | 17 |
| Gynaecological oncologist | 1 | 8 |
| **Area of expertise** |  |  |
| General | 5 | 42 |
| Gynaecologic | 4 | 33 |
| Breast, cervix, colon | 1 | 8 |
| Thoracic, molecular | 1 | 8 |
| Ocular, cytopathology | 1 | 8 |
| **Years of experience** |  |  |
| 0–5 years | 2 | 17 |
| >5 years | 10 | 83 |
| **Institutional affiliation** |  |  |
| Tertiary public hospital | 11 | 92 |
| Private healthcare company | 1 | 8 |

### Current diagnostic context and expressed needs

#### Diagnostic landscape

While H&E staining was universally available and consistently rated high quality, IHC capacity was reported as insufficient for routine diagnosis at 7/10 sites. Molecular testing was entirely absent at 4 sites and limited to HPV DNA testing at 2 (**Figure 1A**). Pathologist availability was limited at most sites, 6/10 had general pathologists only. Satisfaction with pathologist numbers tended to the medium spectrum (median=2.5/5, IQR [1, 3.75]) and most common reported workloads per pathologist were 50-100 to 100-500 cases per week (**Figure 1A**), identical to HIC reference participants.

**Figure 1:**
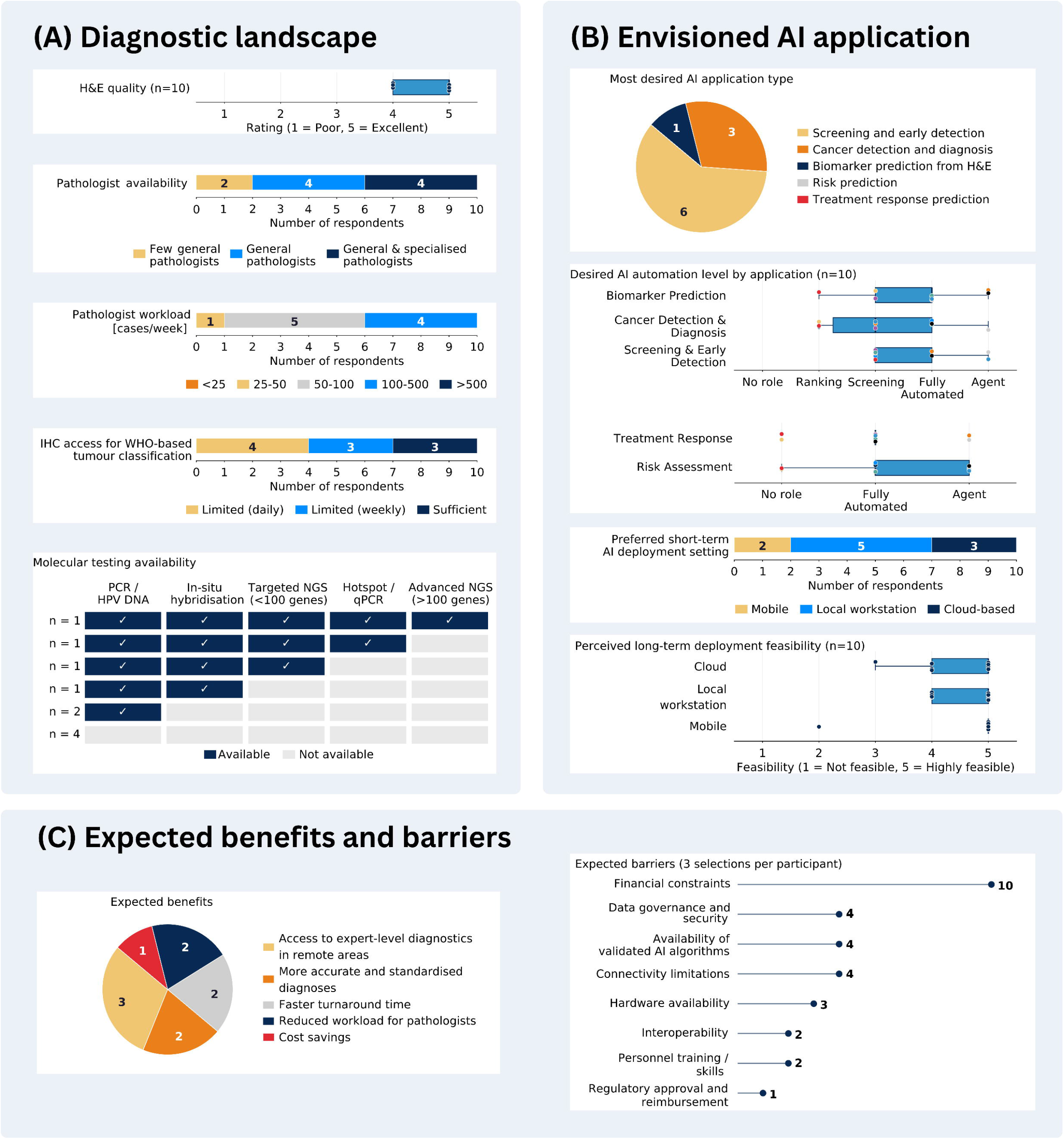
Highlighted survey 1 responses from the 10 LMIC participants. (A) Diagnostic landscape: key characteristics of the current pathology setting, including H&E staining quality, pathologist availability and workload, access to IHC, and molecular testing availability. (B) Envisioned AI application: participants’ preferences for AI application type, desired automation level, and deployment setting. For desired automation level, application types were separated into two categories. For screening and early detection, cancer detection and diagnosis, and biomarker prediction from H&E, where confirmatory pathologist review or IHC/ molecular testing is feasible, five options were offered: no role, ranking (AI orders cases), screening (AI excludes low-risk cases, with confirmatory review of flagged cases), fully automated (AI provides prediction without confirmation), and agentic (AI provides prediction and can initiate downstream actions, such as ordering a test). For risk prediction and treatment response prediction, which represent novel stand-alone biomarker tests without a possible confirmatory step, only three options applied: no role, fully automated, and agentic. Dot colours represent individual respondents. Preferred deployment setting is shown for short (within 1 year) and long-term (5-10 years) horizons across three scenarios: mobile (digitization with smartphone mounted on a microscope, AI run on a smartphone), local workstation (digitization with microscope camera, AI run on a local computer), and cloud-based (digitization with WSI scanner, AI run on a cloud). (C) Expected benefits and barriers to AI implementation in pathology.

#### Envisioned AI application

Cancer screening and early detection was the most frequently selected priority for AI applications, followed by cancer detection and diagnosis, and biomarker prediction from H&E slides (**Figure 1B**). No participant prioritized risk prediction or treatment-response prediction. The median desired level of automation by AI was pre-screening for early detection/screening and diagnosis and full automation for biomarker prediction. All deployment settings were rated as highly feasible in the long-term. In the short-term, local workstation deployment was most preferred, followed by cloud (already implemented at one site), and mobile.

#### Expected benefits and barriers

Expected benefits of AI implementation were broadly distributed across five themes, with improved access to expert-level diagnostics in remote areas selected most frequently (**Figure 1C**). Financial constraints were identified as the dominant anticipated barrier, present in the top three barriers by all participants and rated the most critical barrier by 9/10.

#### Implementation determinants

FGDs identified six implementation determinants (**Figure 2**). AI literacy, evidence generation, governance frameworks, and clinical deployment were perceived as interdependent. Collaboration and funding emerged as cross-cutting factors, both impacting all remaining determinants.

**Figure 2:**
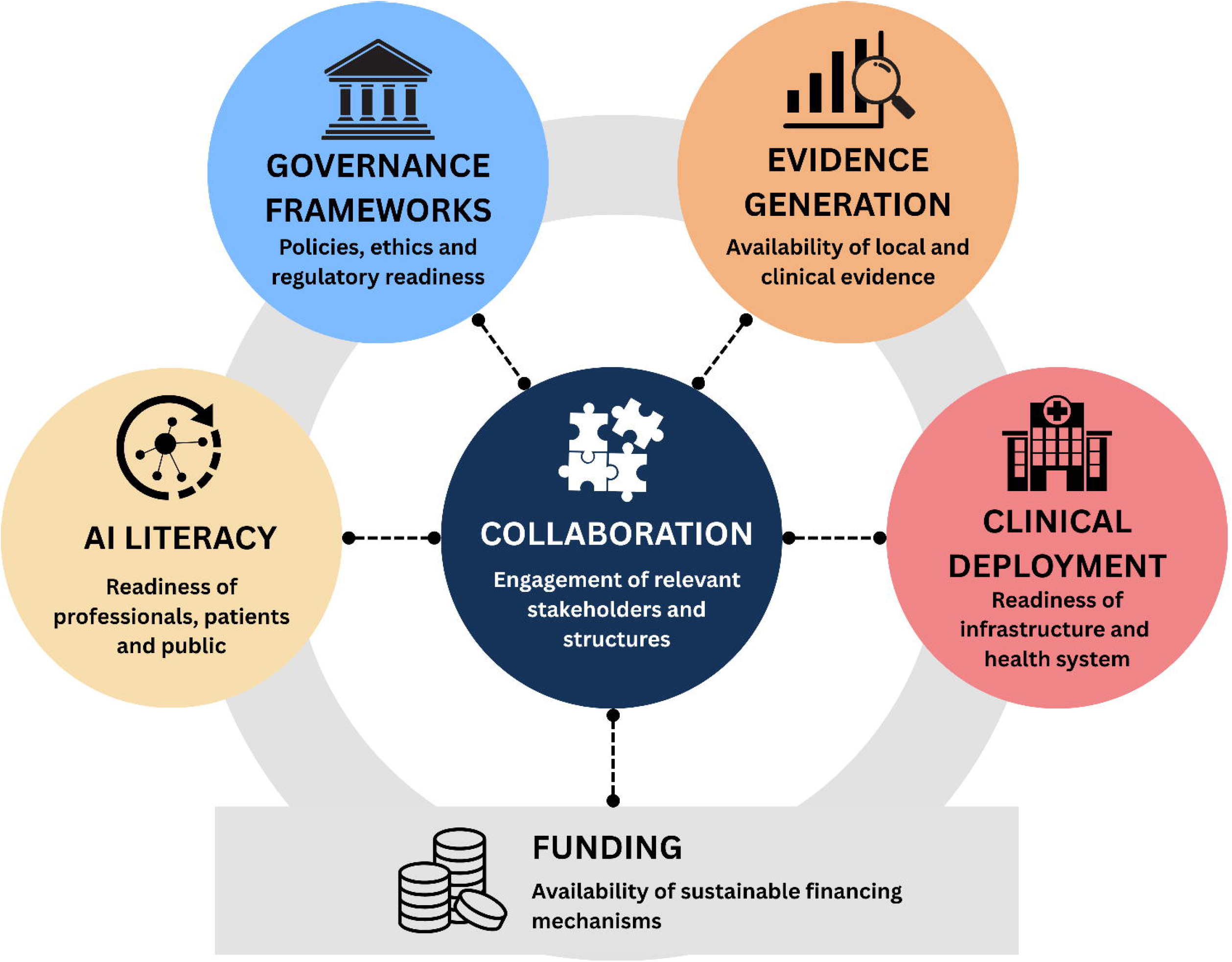
Implementation determinants for AI in LMIC cancer diagnostic settings, identified inductively through focus group discussions. Six interconnected determinants are shown: AI literacy, governance frameworks, evidence generation, and clinical deployment, with collaboration and funding as central mediating factors. The circular arrangement does not imply a sequential or hierarchical order.

#### AI literacy

This determinant captures the social and educational preconditions for AI adoption, the enabling mindset of professionals, patients, and the public to meaningfully engage with AI tools. It comprises three sub-themes: attitudes and acceptability, awareness and sensitisation, and educational integration.

Attitudes and acceptability was the most discussed sub-theme of AI literacy (30 segments, 8/9 participants). For professionals, this centred on trusting AI tools enough to integrate them into practice, moving past perceptions of AI as “*a magic bullet and something that is coming in to replace the human factor in work.*” (Participant (P)07). Participants described acceptance for patients as being mediated through trust in clinicians rather than independent engagement with AI: “*the patient will trust what they will say rather than look at individual results*” (P01). Awareness and sensitisation (14 segments, 6/9 participants) centred on the need to educate both the general public and specific professional groups about existing AI technologies, to avoid knowledge disparities: “*There are people who are very good [with AI], and those who are not aware of it and how it can help.*” (P09). This extended to policymakers, with participants noting that limited AI literacy among decision-makers posed a barrier to the development of relevant policy and regulatory frameworks. Participants linked this directly to educational integration (11 segments, 6/9 participants), emphasizing that “*universities have to embrace AI*” (P07), with some noting existing progress: “*in the university where I do work, now AI is a compulsory course for all university students.*” (P04). Consistent with these themes, in Survey 2, physician attitudes toward AI were mostly rated positive or very positive, while perceived patient attitudes were consistently neutral and general population awareness rated primarily low to moderate (**Figure 3A**).

**Figure 3:**
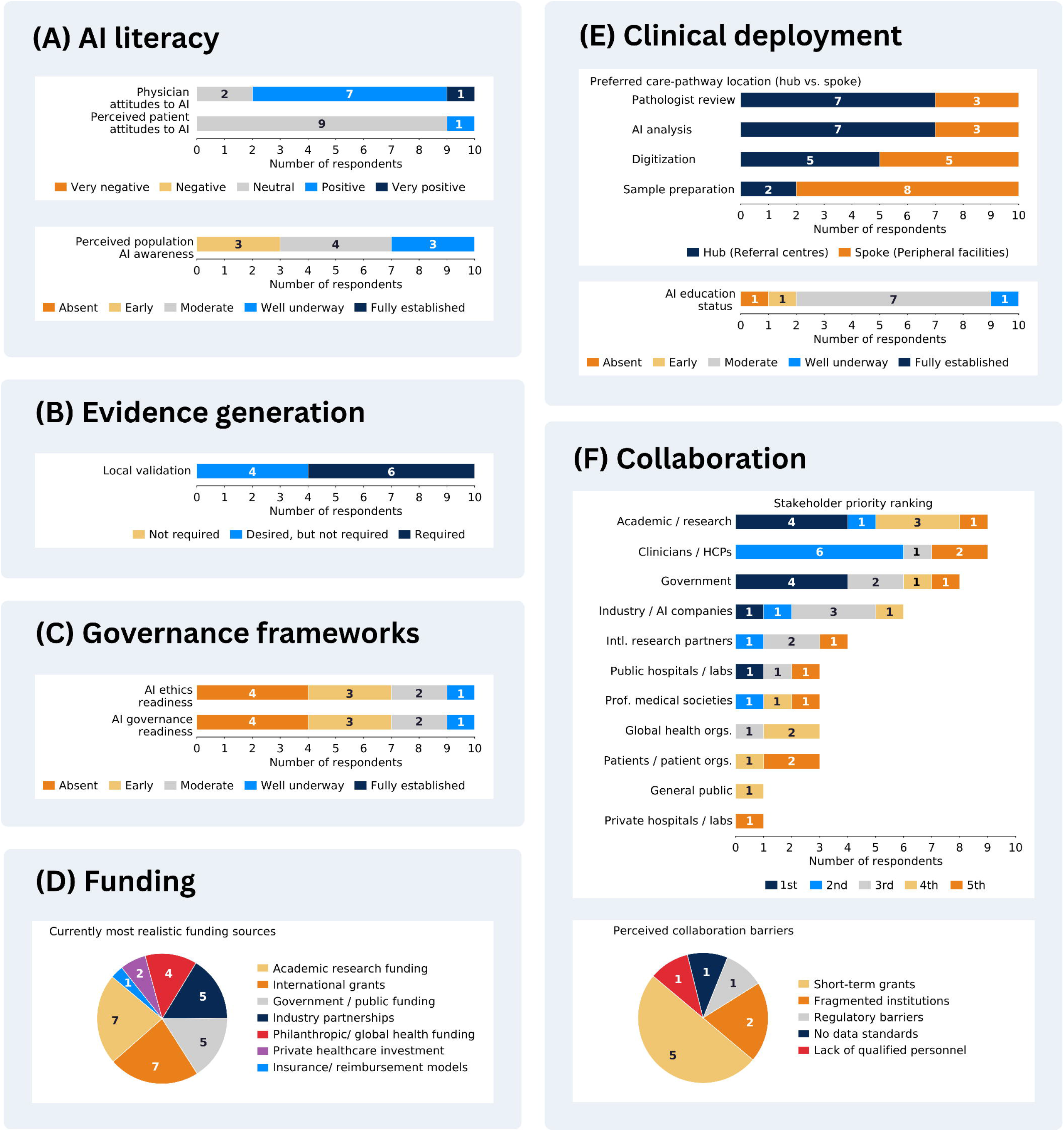
Highlighted survey 2 responses from the 10 LMIC participants, providing quantitative grounding for implementation determinants derived from the focus group discussions. Responses reflect physician perceptions and should be interpreted as such. (A) AI literacy: physician and perceived patient attitudes toward AI, and perceived general population AI awareness. (B) Evidence generation: perceived necessity of local validation of AI algorithms prior to clinical deployment. (C) Governance frameworks: perceived readiness of AI ethics and governance frameworks. (D) Funding: currently most realistic funding sources for AI implementation; up to 3 selections permitted, giving a maximum count of 10 per item. (E) Clinical deployment: preferred care-pathway location for four workflow steps across a hub-and-spoke model, in which hub refers to centralised specialist facilities and spoke to decentralised local facilities; and current status of AI education. (F) Collaboration: stakeholder priority ranking among participants’ top 5 most important stakeholders for AI implementation (colours indicate rank position); and perceived primary barrier to collaboration (single choice).

#### Evidence Generation

This determinant captures the perceived need for research demonstrating AI’s clinical benefit, encompassing two sub-themes: research and pilot studies and local validation.

Research and pilot studies (9 segments, 4/9 participants) were viewed as a precursor to securing funding, by building a concrete case for AI’s benefit: “*when you demonstrate the utility, then you can put this on the table to show that this has a benefit … on the patient. And then you can have reimbursement for those applications.*” (P03). University hospitals were described as central actors in generating this evidence, with participants citing examples such as cervicography-based cervical cancer screening and AI-assisted assessment of biomarkers (ER, PR, HER2). Local validation (3 segments, 2/9 participants) was considered essential to ensure AI outputs align with local population characteristics and diagnostic workflows, given that tools developed in high-income settings may not transfer without adaptation. One participant warned that without it, “*people might lose interest in this AI-assisted cancer diagnosis.*” (P04). Survey 2 showed that most respondents considered local validation studies required, with the remainder viewing them as desired but not strictly necessary (**Figure 3B**).

#### Governance frameworks

This determinant captures perceived needs and gaps in policy, regulation, and ethical frameworks governing AI use in clinical practice, comprising four sub-themes: development of guidelines, development of policies/legislation, ethics/consent and regulatory readiness/gaps.

Regulatory readiness/gaps (14 segments, 7/9 participants) was the most frequently discussed sub-theme, capturing a perceived mismatch between the pace of AI adoption and that of regulatory development: “*the regulators actually are a bit behind the practitioners in terms of appreciating the necessity to move much faster*.” (P05). The majority of participants raised the need for development of national AI strategies, data protection legislation, and policies governing AI use in clinical practice (13 segments, 6/9 participants), alongside clinician- and professional society-led consensus on acceptable guidelines, such as “*consensus on guidelines and regulations as to what is acceptable in the histopathology report. How much of AI assisted work can be accepted within the report.*” (2 segments, 1/9 participants). In Survey 2, AI governance and ethics frameworks were most rated as absent or early-stage, with no participant rating either as fully established (**Figure 3C**), including HIC participants. Ethics/consent (13 segments, 6/9 participants) centred on unresolved accountability concerns that participants felt could delay adoption: “*If something got read in the AI project and there was a dilemma, who is liable for this? There are no laws as yet.*” (P07). The need for healthcare providers to be trained in ethical AI use was also emphasised.

#### Clinical deployment

This determinant captures perceived requirements for deploying AI tools in clinical routine, comprising six sub-themes: infrastructure, deployment strategy, envisioned role of AI, equity of access and the current healthcare landscape, pathologist workforce, and training of healthcare professionals.

Infrastructure was the most frequently discussed sub-theme (38 segments, 9/9 participants), spanning digitization and scanning equipment, connectivity, hardware costs, and broader infrastructure readiness. Survey 1 identified available WSI scanners at 8/10 LMIC sites and GPUs at 4/10. Considerable heterogeneity in, and discordance between, AI and clinical infrastructure levels were observed (**Figure 4**). Settings with relatively high AI infrastructure scores and limited IHC, molecular testing, and pathologist availability, represent promising candidates for leapfrogging. The FGD participants described highly variable situations, some reporting adequate resources (“*even some people have small scanner[s]*” (P09)), and others citing critical gaps, with one noting that building a digital pathology workflow is less costly than replicating a good quality analogue system: “*the cost is way higher to go analog than to go digital*” (P08), highlighting also an economic rationale for leapfrogging.

**Figure 4:**
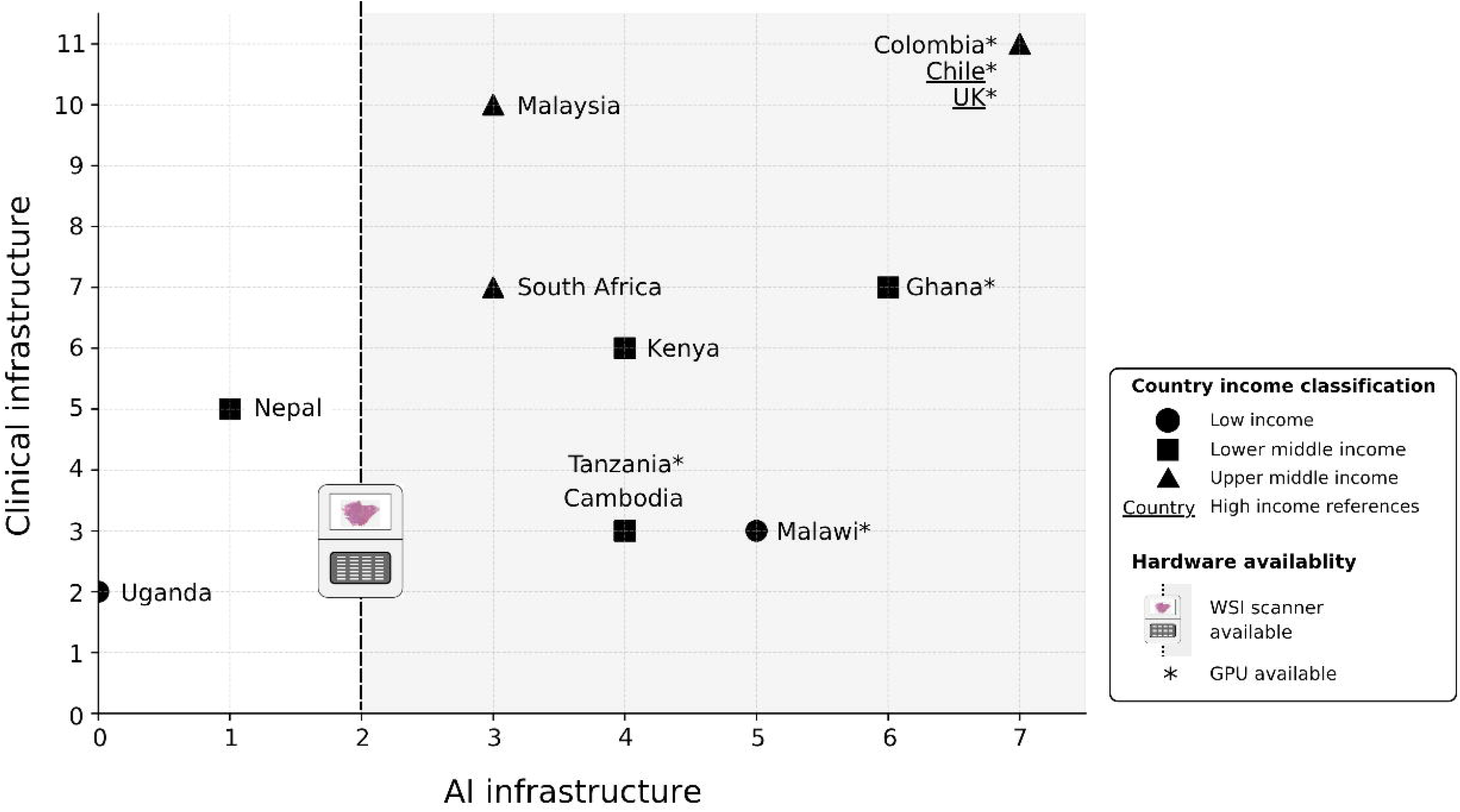
Level of clinical and AI infrastructure scores across participants. AI infrastructure (x-axis, maximum score 7) and clinical infrastructure (y-axis, maximum score 11) were assessed using study-specific scoring heuristics applied to Survey 1 responses. Each data point represents a single participant; country labels are provided for orientation only and should not be interpreted as nationally representative infrastructure scores. The dashed vertical line separates participants reporting no WSI scanner available (left) from those reporting at least a low-throughput WSI scanner (right); GPU availability is indicated by *. HIC participants (Chile, UK; <u>underlined</u>) are included as illustrative reference points.

Deployment strategy (20 segments, 8/9 participants) discussions pointed toward integrating AI into existing healthcare pathways in a phased manner, beginning at better-resourced tertiary or university hospitals before broader scale-up: “*a good implementation would be one that integrates… the county healthcare system [and] would reach even the very remote places.*” (P08). Opinions differed on the optimal model, some favoured decentralized AI at secondary facilities closer to patients, while others advocated for centralized digital pathology hubs connected through telepathology networks, with peripheral facilities serving as sample collection points. Survey 2 quantified this hub-and-spoke framing: sample preparation was envisioned at the spoke by most respondents, while AI analysis and reporting were envisioned primarily at the hub, reflecting a pattern of progressive centralization (**Figure 3E**).

Equity of access discussions (23 segments, 8/9 participants) identified geographic and financial barriers to care, with AI viewed as a potential equalizer extending diagnostic access to underserved regions (“*it’s possible to bridge those distance gaps or human resource gaps using tech. And I think AI would come in there.*” (P08)), though participants cautioned against solutions that might “*duplicate the same limitation in a new setting.*” (P07).

Pathologist workforce (14 segments, 8/9 participants) and the envisioned role of AI (12 segments, 7/9 participants) reflected the Survey 1 results, highlighting existing workforce shortages and AI envisioned as a collaborative tool augmenting rather than replacing pathologist judgment. Training of healthcare professionals (20 segments, 7/9 participants) emphasized the need to embed AI education structurally in both initial training and ongoing professional development. Consistent with this, in Survey 2, current AI education for healthcare providers was mostly rated as no more than moderately integrated (**Figure 3E**). The most frequently selected priority topics for training were AI implementation in clinical practice (6/10) and basic AI concepts (5/10), and the most preferred formats were integration into pathology residency training (9/10) and short in-person courses (7/10).

## Funding

This determinant captures perceived sources of funding for AI implementation in cancer diagnostics, comprising four sub-themes: government/public funding, (international) research funding, reimbursement and insurance, and private sector/public-private partnerships.

Government and public funding was the most frequently discussed sub-theme (17 segments, 7/9 participants), centring on the constraints posed by limited health budgets and the need for greater government investment. Local or international research funding (6 segments, 5/9 participants) was viewed as a viable source of both funding and equipment. Reimbursement and insurance (10 segments, 4/9 participants) centred on the current status of insurance frameworks and their potential role in sustaining AI deployment. Expanding public health insurance coverage was seen as the most plausible pathway to financial viability of AI deployment, though exclusion from current reimbursement structures was identified as a barrier: “*the payment for the services, it’s not included AI… that’s one point why we do not have AI already performing*” (P03). Private sector and public-private partnerships (10 segments, 5/9 participants) were seen as a route to offsetting high infrastructure costs and accessing advanced technology: “*it’s possible to vet private systems and make a partnership.*” (P08). In Survey 2, however, academic research funding and international grants were selected as the most realistic funding sources, ahead of government and public funding (**Figure 3D**).

### Collaboration

This determinant captures the stakeholders and collaborative structures, the relational layer needed to advance all preceding determinants.

### Key stakeholders

Government was the most prominently discussed stakeholder (50 segments, 8/9 participants), with participants emphasizing the roles of ministries in funding, regulation, and national strategy. Academic and research institutions (37 segments, 8/9 participants) and public healthcare systems (35 segments, 8/9 participants) were similarly prominent, with universities and government hospitals identified as the primary sites for evidence generation, training, and deployment. Clinicians and healthcare providers (31 segments, 9/9 participants) were highlighted as essential end-users, particularly pathologists, whose role in validating AI outputs and integrating findings into multidisciplinary teams was consistently emphasized. The private sector, including industry/AI vendors (10 segments, 6/9 participants) and private healthcare companies (15 segments, 5/9 participants), was discussed as a key funding and implementation partner including through direct provision of equipment and leveraging existing private diagnostic infrastructure. Patients and the general public (12 and 9 segments respectively) and professional medical societies (9 segments, 6/9 participants) were recognized as important but less frequently discussed stakeholders. In Survey 2, government and academic/research groups were most frequently ranked as the top-priority stakeholder, with clinicians and industry appearing consistently across the top five ranks (**Figure 3F**), consistent with the FGD appearance frequency.

### Collaboration structures

Participants framed collaboration as an urgent necessity, noting that current efforts remain fragmented. Sustainability of partner-driven initiatives was also raised as a concern, with one participant cautioning that projects “*might work now, but in the long term, perhaps sustainability can be an issue.*” (P01). To address these challenges, participants proposed several collaborative structures. Research collaboration, both national and international, was the most frequently discussed collaboration type (11 segments, 5/9 participants), encompassing co-creation of locally relevant solutions, multi-institutional and potentially international partnerships: “*it can even go beyond boundaries… this can bring sets of the global-local alliance*” (P04). Public-private partnerships (8 segments, 4/9 participants) were highlighted as a viable model for offsetting hardware and infrastructure costs: “*we have to get together with government, and professional bodies and vendors… it’s got to be mutually beneficial.*” (P05). Advocacy and lobbying (7 segments, 3/9 participants) centred on the need for unified scientific and professional voices to influence policy and secure funding. Clinical-university collaboration (4 segments, 4/9 participants) was described as foundational for generating and translating evidence into clinical practice. In Survey 2, the collaboration types rated most important were also most frequently absent, specifically government-academic, public-private partnerships, and international research collaboration. Dependence on short-term grants or donor cycles was the most frequently reported barrier to sustainable collaboration (**Figure 3F**).

## Discussion

The findings of this study show that readiness for implementation of AI in cancer diagnostics in LMIC settings is highly heterogeneous, cannot be inferred from country income group alone, and is constrained across all dimensions by financial barriers and lack of essential collaborations.

Clinical and AI infrastructure varied substantially: some settings showed relatively high AI infrastructure alongside limited clinical capacity, indicating potential for leapfrogging (8), while others showed the inverse. This finding suggests that income classification, while a common organising framework in global AI readiness assessments (24), is an insufficient proxy for deployment readiness at the institutional level, underscoring the need for context-specific feasibility assessment before AI implementation.

The predominant clinical infrastructure gaps were insufficient IHC capacity, absent or minimal molecular testing, and shortage of pathologists. High-quality H&E staining was universally available, providing an essential element of H&E-based AI-assisted cancer screening and diagnosis to offset lack of pathologist capacity, and H&E-based biomarker prediction to improve allocation of limited IHC and molecular tests. The availability of mainly only low-throughput scanners will, however, limit the scale of the impact: digitising 1-6 slides per batch at a few minutes per slide, such devices can process up to several hundred slides per day under continuous operation, which may suffice at the facility level but becomes a bottleneck when few devices serve an entire country or region. At the same time, conventional high-throughput scanning infrastructure represents a cost barrier, estimated at over $1.6 million annually at a large HIC centre (25). Intermediate digitisation solutions, from open-source microscopes (26), microscope cameras (27) to low-throughput scanners, may therefore represent more realistic near-term entry points for many LMIC settings. Limited GPU availability at most LMIC settings points to two complementary directions: investment in GPU capacity, as well as development of algorithms with reduced computational requirements (28,29).

Beyond infrastructure, six implementation determinants emerged from the qualitative phase: AI literacy, governance frameworks, evidence generation, clinical deployment, funding and collaboration. They are broadly consistent with HIC-derived frameworks such as FAIIR-H (30) and HAIRA (31), suggesting core implementation determinants may generalize across settings. However, several LMIC-specific nuances emerged. AI literacy among clinicians was identified as a critical precondition, with clinician-perceived gatekeeping of patient acceptability, a dimension addressed primarily at the organisational level in existing implementation frameworks. Considerable heterogeneity was observed in application types and desired automation level across participants. This points to a spectrum of adopter attitudes that AI implementation strategies will need to accommodate, consistent with evidence of varying adoption profiles in digital health (13,32). Governance and ethics frameworks were nascent across most settings, consistent with global assessments documenting a sustained HIC advantage in AI governance instruments (24,33). Beyond the structural absence or incompleteness of frameworks participants identified limited AI literacy among policymakers, a nuance not captured in frameworks assuming technically informed governance actors (30,31). All participants considered local validation necessary or desirable before AI deployment, reflecting generalization concerns documented in prior studies (34). Responsibility for generating this evidence was discussed primarily in relation to local clinical-academic teams. We argue that this responsibility should be shared with AI developers and industry partners, together producing context-specific validation data for LMIC settings, and ensuring equitable benefit-sharing when patient data from these settings inform commercial model development. For clinical deployment, participants envisioned a phased rollout beginning at tertiary centres, with AI analysis centralised at hubs and sample preparation at peripheral sites, consistent with proposed hub-and-spoke strategies for digital pathology scale-up in LMICs (35). Such models offer a pragmatic pathway for scale-up, but realising their potential requires deliberate planning to ensure implementation reaches peripheral and underserved settings to reduce rather than reinforce existing diagnostic disparities.

Financial constraints emerged not as a domain-specific barrier but as the structural bottleneck for all other implementation determinants, surfacing across hardware acquisition and maintenance, regulatory development, educational integration, evidence generation, and collaboration sustainability. Government funding was most frequently mentioned in FGDs as the appropriate long-term source, yet academic research and international grants were most frequently identified as realistically available near-term. Grant and donor funding, however, typically covers capital costs rather than the long-term costs of upkeep and repair, leaving infrastructure at risk of becoming unused or inoperable if local capacity to maintain and operate AI tools is not built in parallel. This reflects the structural reality that government health expenditure in LMICs remains well below recommended thresholds, with out-of-pocket payments the dominant financing source in many settings (36), mirroring well-documented tensions between aspirational and operational funding models in global health (37). A parallel gap is evident in collaboration: the types of partnership considered most essential, government-academic, academic-industry, and international research, were also those most frequently identified as currently absent, with dependence on short-term grants the dominant barrier to initiating and sustaining collaboration (38,39). Overall, these findings suggest that addressing individual implementation determinants is unlikely to achieve durable impact without sustainable, institutionally embedded financing models.

This study has several strengths and limitations. The sequential mixed-methods design enabled a structured, iterative approach, first characterising the diagnostic landscape and expressed needs, then inductively identifying implementation determinants, and finally quantifying them, providing a methodological basis for larger-scale assessment. Purposive recruitment achieved broad geographic representation across twelve countries and four regions, with each participant representing a distinct national context, while double-coded qualitative analysis with iterative codebook refinement strengthened the credibility of the qualitative findings. However, the moderate sample size (N=10 LMIC) limits generalisability, and findings should be considered exploratory rather than representative. Recruitment through professional networks and publication authorship lists likely favoured academically engaged, AI-familiar clinicians from tertiary or university hospitals, potentially overestimating implementation readiness and underestimating barriers in less-resourced settings. The study was further limited to physicians, excluding perspectives from policymakers, laboratory technicians, IT staff, hospital managers, and patients, whose roles in AI implementation readiness may be substantial and warrant inclusion in future work. All data were self-reported by physicians, which may introduce inaccuracies for domains outside their direct purview, such as digital infrastructure, AI literacy levels and governance readiness. Country-level findings reflect the perspectives of single participants and should not be interpreted as nationally representative, and the self-reported readiness assessments and custom infrastructure scoring heuristic were not independently validated.

Taken together, this study provides preliminary empirical evidence from a geographically diverse LMIC cancer-diagnostic sample for the determinants shaping AI implementation readiness. The six implementation determinants extend beyond existing HIC-derived frameworks, with financial constraints as the universal cross-cutting barrier and collaboration as a crucial mechanism through which each factor can be advanced.

Future work should prioritise larger, more diverse samples including primary and secondary facility staff, direct assessment of patient attitudes toward AI, longitudinal evaluation of how readiness evolves as regulatory frameworks and financing models mature, and per-country, cancer-type-specific analyses of AI application priorities. These steps would allow translation of the implementation determinants identified here into actionable guidance for AI-enabled cancer diagnostics in resource-limited settings.

## Conclusions

This study empirically identified six implementation determinants shaping readiness for AI adoption in cancer diagnostics in LMICs, providing a preliminary basis for larger-scale, representative assessment. AI may offer a leapfrogging opportunity in some contexts, by allocating and supporting limited diagnostic capacity. Realising the potential of AI requires sustained collaboration and financing embedded within health system budgets rather than relying on short-term donor funding, to invest in health care provider education, technical and clinical infrastructure.

### Declaration of competing interest

T.B and N.H. received research funding from the Dutch Cancer Society and are supported by the Hanarth Fund, unrelated to the present study. V.K. is a participant in several patent applications on the assessment of cancer biomarkers by digital pathology. T.B., V.K. and N.H. are participants on a patent application on multimodal deep learning for the prediction of recurrence risk in endometrial cancer patients. V.K. reports sponsored research agreements with Roche and IAG, honoraria or consulting fees from Takeda and Roche, and participation as an invited speaker for Sharing Progress in Cancer Care and Indica Labs, all unrelated to the present study. The other authors declare no competing interests.

### Data availability statement

Survey instruments, results, Survey 1 guide, the focus group discussion guide and qualitative codebook are provided as supplementary material. Anonymised focus group transcripts may be made available on reasonable request to the corresponding author, subject to data privacy review and consistency with participant consent.

### Author contributions

Lydia A Schoenpflug: Conceptualization, Data curation, Formal analysis, Investigation, Methodology, Project administration, Software, Validation, Visualization, Writing - Original Draft, Writing - Review & Editing; Nikki J van den Berg: Conceptualization, Data curation, Formal analysis, Investigation, Methodology, Project administration, Writing - Review & Editing; Serey Vathana Chhut: Writing - Review & Editing; Elaine YL Leung: Writing - Review & Editing; Lai-Meng Looi: Writing - Review & Editing; Robert Lukande: Writing - Review & Editing; Samuel James Meja: Writing - Review & Editing; Rafael Parra-Medina: Writing - Review & Editing; Tilak Pathak: Writing - Review & Editing; Jeremia J Pyuza: Writing - Review & Editing; Rubina Razack: Writing - Review & Editing; Augustine Tawiah: Writing - Review & Editing; Maureen W Waithaka: Writing - Review & Editing; Pablo Zorroquiain: Writing - Review & Editing; Jurriaan Barkey Wolf: Writing - Review & Editing; Marcus J Rijken: Writing - Review & Editing;Gernot Jundt: Writing - Review & Editing; Maxime W Lafarge: Supervision, Writing - Review & Editing; Tjalling Bosse: Conceptualization, Funding acquisition, Resources, Supervision, Writing - Review & Editing; Viktor H Koelzer: Conceptualization, Funding acquisition, Resources, Supervision, Writing - Review & Editing; Nanda Horeweg: Conceptualization, Data curation, Investigation, Methodology, Project administration, Supervision, Validation, Writing - Review & Editing

### Funding statement

This study received no specific grant funding from public, commercial, or not-for-profit funding agencies. The work was supported by core institutional funding from the University of Basel.

## Supporting information

Supplementary File 1: GRAMMS checklist

Supplementary File 2: CHERRIES checklist

Supplementary File 3: COREQ checklist

Supplementary File 4: Survey 1 instrument

Supplementary File 5: Survey 1 guide

Supplementary File 6: Survey 1 responses

Supplementary File 7: Survey 2 instrument

Supplementary File 8: Survey 2 responses

Supplementary File 9: Focus group discussion guide

Supplementary File 9: Focus group discussion code book

## Acknowledgements

The authors thank Mariam Gachechiladze, University Hospital Zürich, for reviewing the Survey 1 instrument and providing expert guidance on their relevance to LMIC cancer-diagnostic contexts. We are grateful to all participants for their time and willingness to share their experiences and perspectives across multiple study phases.

## Supplementary

● Supplementary File 1: GRAMMS checklist

● Supplementary File 2: CHERRIES checklist

● Supplementary File 3: COREQ checklist

● Supplementary File 4: Survey 1 instrument

● Supplementary File 5: Survey 1 guide

● Supplementary File 6: Survey 1 responses

● Supplementary File 7: Survey 2 instrument

● Supplementary File 8: Survey 2 responses

● Supplementary File 9: FGD guide

● Supplementary File 10: FGD code book

## Notes

### Author Declarations

Ethics Committee of Northwest and Central Switzerland (EKNZ) waived ethical approval for this work.

### Summary of Updates

Authors updated; Methods updated for completeness and clarity; Deeper consideration of impact of funding limitations in Discussion.

