## Supplementary File 1: GRAMMS checklist for "Leapfrogging cancer diagnostics with AI: a mixed-methods assessment of implementation determinants in low- and middle-income countries"

Good Reporting of A Mixed Methods Study (GRAMMS) checklist

Developed from:

O'Cathain A, Murphy E, Nicholl J. The quality of mixed methods studies in health services research. J Health Serv Res Policy. 2008;13: 92-98.

| **Checklist item** | **Guide Questions/Description** | **Response** |
| --- | --- | --- |
| Justification | Describe the justification for using a mixed methods approach to the research question | Infrastructure-related readiness factors are directly observable and appropriately assessed through structured survey. Non-infrastructure factors, including governance, funding structures, stakeholder roles, and collaboration are not well characterised in LMIC cancer-diagnostic contexts and could not be pre-specified from existing literature. Qualitative exploration was required to identify which factors were locally relevant before they could be systematically measured. A mixed methods approach enabled both direct infrastructure measurement and the discovery and subsequent quantification of non-infrastructure factors within the same study population. |
| Design | Describe the design in terms of the purpose, priority and sequence of methods | An exploratory sequential design was used (Quantitative, Qualitative, Quantitative), following an instrument-development logic. Survey 1 established the infrastructure baseline; FGDs identified implementation factors relevant to this population; Survey 2 quantified those factors. Each phase directly informed the design of the next. |
| Methods | Describe each method in terms of sampling, data collection and analysis | Survey 1: 70-item structured web-based questionnaire, interviewer-guided, purposive sample of 12 LMIC clinicians, analysed descriptively. FGDs: nine participants across three online sessions, hybrid inductive-deductive thematic analysis, independent double-coding. Survey 2: 18-item self-administered web-based questionnaire, same sample, analysed descriptively. |
| Integration | Describe where integration has occurred, how it has occurred and who has participated in it | Integration occurred at three points: Survey 1 findings informed the FGD guide; FGD themes were operationalised into Survey 2 by LAS and NH through investigator consensus; all three data sources were compared during interpretation through narrative triangulation by LAS and NH. |
| Limitations | Describe any limitation of one method associated with the present of the other method | The sequential design introduced a seven-month gap between Survey 1 and Survey 2, limiting temporal comparability. Prior survey exposure may have implicitly framed FGD discussions toward infrastructure topics. Survey 2's self-administered format, adopted given participants' familiarity with study terminology, may have introduced response heterogeneity on nuanced items. |
| Insights | Describe any insights gained from mixing or integrating methods | Integration produced findings not identifiable from any single method alone. The divergence between government funding as the most discussed FGD topic and the third-ranked realistic near-term source in Survey 2 was only visible through cross-phase comparison. The provider-patient attitude gap identified in Survey 2 was interpretable only through the FGD finding that patients rely on clinician endorsement. The finding that most-needed collaboration types are also most currently absent required FGDs to identify the relevant categories and Survey 2 to quantify their absence. |
