## Supplementary File 2: CHERRIES checklist for "Leapfrogging cancer diagnostics with AI: a mixed-methods assessment of implementation determinants in low- and middle-income countries"

Checklist for Reporting Results of Internet E-Surveys (CHERRIES)

This checklist has been modified from Eysenbach G. Improving the quality of Web surveys: the Checklist for Reporting Results of Internet E-Surveys (CHERRIES). J Med Internet Res. 2004 Sep 29;6(3):e34 [erratum in J Med Internet Res. 2012; 14(1): e8.]. Article available at https://www.jmir.org/2004/3/e34/; erratum available https://www.jmir.org/2012/1/e8/. Copyright ©Gunther Eysenbach. Originally published in the Journal of Medical Internet Research, 29.9.2004 and 04.01.2012.

This is an open-access article distributed under the terms of the Creative Commons Attribution License (https://creativecommons.org/licenses/by/2.0/), which permits unrestricted use, distribution, and reproduction in any medium, provided the original work, first published in the Journal of Medical Internet Research, is properly cited.

| **Checklist item** | **Guide Questions/Description** | **Response** |
| --- | --- | --- |
| Describe survey design | Describe target population, sample frame. Is the sample a convenience sample? (In “open” surveys this is most likely.) | Both surveys were closed, cross-sectional, web-based questionnaires administered via Google Forms to a purposively recruited sample of clinicians involved in cancer diagnostics across LMIC settings. Survey 1 assessed clinical and AI infrastructure, AI application preferences, and implementation barriers. Survey 2 assessed perceived implementation readiness across five FGD-derived domains. The sample was purposive, not a convenience sample; participants were individually identified and contacted by the research team. |
| IRB approval | Mention whether the study has been approved by an IRB. | The study was exempted from formal ethics review by the Ethics Committee Northwest and Central Switzerland (EKNZ). |
| Informed consent | Describe the informed consent process. Where were the participants told the length of time of the survey, which data were stored and where and for how long, who the investigator was, and the purpose of the study? | Informed electronic consent was obtained via Google Forms. Participants were informed of the study objectives, the research team's institutional affiliation, the voluntary nature of participation, data storage procedures, and their right to request data deletion. Participants indicated their preference regarding future data use (deletion, recontact, or unrestricted future use). The approximate duration of Survey 1 (approximately one hour) and Survey 2 (approximately 15 minutes) was communicated to participants prior to each session. |
| Data protection | If any personal information was collected or stored, describe what mechanisms were used to protect unauthorized access. | Data were stored on an institutional server with restricted access. |
| Development and testing | State how the survey was developed, including whether the usability and technical functionality of the electronic questionnaire had been tested before fielding the questionnaire. | Both surveys were developed iteratively and reviewed by a regional and methodological expert with LMIC experience for clarity and relevance. Survey 1 was piloted with one participant prior to data collection to assess usability, comprehension, and completion time; identified issues were resolved before fielding. Survey 2 was reviewed by the investigator team but was not separately piloted given its shorter length and the participants' prior familiarity with study terminology from Survey 1 and the FGDs. |
| Open survey versus closed survey | An “open survey” is a survey open for each visitor of a site, while a closed survey is only open to a sample which the investigator knows (password-protected survey). | Both surveys were closed, accessible only to individually invited participants via a direct link shared by the research team. The surveys were not publicly advertised or open to general visitors. |
| Contact mode | Indicate whether or not the initial contact with the potential participants was made on the Internet. (Investigators may also send out questionnaires by mail and allow for Web-based data entry.) | Initial contact was made via email, not via the internet survey platform itself. |
| Advertising the survey | How/where was the survey announced or advertised? Some examples are offline media (newspapers), or online (mailing lists – If yes, which ones?) or banner ads (Where were these banner ads posted and what did they look like?). It is important to know the wording of the announcement as it will heavily influence who chooses to participate. Ideally the survey announcement should be published as an appendix. | Neither survey was advertised publicly. Recruitment was conducted through direct, individualised email invitation to identified potential participants. No banner ads, mailing lists, or social media were used. |
| Web/E-mail | State the type of e-survey (eg, one posted on a Web site, or one sent out through e-mail). If it is an e-mail survey, were the responses entered manually into a database, or was there an automatic method for capturing responses? | Both surveys were web-based questionnaires hosted on Google Forms. Links were distributed via email. Responses were captured automatically by Google Forms and exported for analysis; no manual data entry was required. |
| Context | Describe the Web site (for mailing list/newsgroup) in which the survey was posted. What is the Web site about, who is visiting it, what are visitors normally looking for? Discuss to what degree the content of the Web site could pre-select the sample or influence the results. For example, a survey about vaccination on a anti-immunization Web site will have different results from a Web survey conducted on a government Web site | Surveys were not posted on a public website. Links were shared directly via email to identified participants. The Google Forms platform is a general-purpose survey tool with no thematic content that could pre-select or influence the sample. |
| Mandatory/voluntary | Was it a mandatory survey to be filled in by every visitor who wanted to enter the Web site, or was it a voluntary survey? | Participation was entirely voluntary. No access restriction or incentive was tied to survey completion. |
| Incentives | Were any incentives offered (eg, monetary, prizes, or non-monetary incentives such as an offer to provide the survey results)? | Yes, preliminary access to the survey results and review of the study manuscript. |
| Time/Date | In what timeframe were the data collected? | Survey 1: October-November 2025. Survey 2: June 2026. |
| Randomization of items or questionnaires | To prevent biases items can be randomized or alternated. | No randomisation of items or questionnaire order was used in either survey. |
| Adaptive questioning | Use adaptive questioning (certain items, or only conditionally displayed based on responses to other items) to reduce number and complexity of the questions. | No adaptive questioning or skip logic was used in either survey. Both surveys used a fixed, linear item sequence. |
| Number of Items | What was the number of questionnaire items per page? The number of items is an important factor for the completion rate. | Survey 1: 70 items across 7 sections. Survey 2: 18 items across 5 sections. |
| Number of screens (pages) | Over how many pages was the questionnaire distributed? The number of items is an important factor for the completion rate. | Survey 1: 7 screens, Survey 2: 5 screens |
| Completeness check | It is technically possible to do consistency or completeness checks before the questionnaire is submitted. Was this done, and if “yes”, how (usually JAVAScript)? An alternative is to check for completeness after the questionnaire has been submitted (and highlight mandatory items). If this has been done, it should be reported. All items should provide a non-response option such as “not applicable” or “rather not say”, and selection of one response option should be enforced. | All items in both surveys were set as mandatory in Google Forms, preventing submission of incomplete responses. No item offered a "not applicable" or "prefer not to say" option; selection of a response was enforced for all items. No JavaScript-based consistency checks were applied; completeness was enforced at the point of submission. |
| Review step | State whether respondents were able to review and change their answers (eg, through a Back button or a Review step which displays a summary of the responses and asks the respondents if they are correct). | Google Forms allows participants to scroll back and amend responses at any point before final submission. A review of all responses was possible prior to submitting the form. |
| Unique site visitor | If you provide view rates or participation rates, you need to define how you determined a unique visitor. There are different techniques available, based on IP addresses or cookies or both. | Not applicable. Both surveys were closed and distributed via individual email links to identified participants. Unique participation was ensured through the direct invitation model rather than through technical tracking mechanisms. |
| View rate (Ratio of unique survey visitors/unique site visitors) | Requires counting unique visitors to the first page of the survey, divided by the number of unique site visitors (not page views!). It is not unusual to have view rates of less than 0.1 % if the survey is voluntary. | Not applicable. Surveys were not hosted on a public website; no site visitor data were recorded. |
| Participation rate (Ratio of unique visitors who agreed to participate/unique first survey page visitors) | Count the unique number of people who filled in the first survey page (or agreed to participate, for example by checking a checkbox), divided by visitors who visit the first page of the survey (or the informed consents page, if present). This can also be called “recruitment” rate. | Of 15 individuals contacted, 13 completed Survey 1 (87%). All 12 analysed participants subsequently completed Survey 2 (100% of the analysed sample). |
| Completion rate (Ratio of users who finished the survey/users who agreed to participate) | The number of people submitting the last questionnaire page, divided by the number of people who agreed to participate (or submitted the first survey page). This is only relevant if there is a separate “informed consent” page or if the survey goes over several pages. This is a measure for attrition. Note that “completion” can involve leaving questionnaire items blank. This is not a measure for how completely questionnaires were filled in. (If you need a measure for this, use the word “completeness rate”.) | 100% for both surveys among participants who began the form, as mandatory response settings prevented partial submission. |
| Cookies used | Indicate whether cookies were used to assign a unique user identifier to each client computer. If so, mention the page on which the cookie was set and read, and how long the cookie was valid. Were duplicate entries avoided by preventing users access to the survey twice; or were duplicate database entries having the same user ID eliminated before analysis? In the latter case, which entries were kept for analysis (eg, the first entry or the most recent)? | Google Forms does not set tracking cookies for survey response purposes. No cookies were used to track or identify participants. |
| IP check | Indicate whether the IP address of the client computer was used to identify potential duplicate entries from the same user. If so, mention the period of time for which no two entries from the same IP address were allowed (eg, 24 hours). Were duplicate entries avoided by preventing users with the same IP address access to the survey twice; or were duplicate database entries having the same IP address within a given period of time eliminated before analysis? If the latter, which entries were kept for analysis (eg, the first entry or the most recent)? | No IP address tracking was used. Duplicate entries were not a concern given the closed, individually invited design and the small, known sample. |
| Log file analysis | Indicate whether other techniques to analyze the log file for identification of multiple entries were used. If so, please describe. | No log file analysis was conducted. |
| Registration | In “closed” (non-open) surveys, users need to login first and it is easier to prevent duplicate entries from the same user. Describe how this was done. For example, was the survey never displayed a second time once the user had filled it in, or was the username stored together with the survey results and later eliminated? If the latter, which entries were kept for analysis (eg, the first entry or the most recent)? | Both surveys were closed. Links were distributed individually via email. No formal login or registration system was used; access control relied on individual distribution of unique links to identified participants rather than a technical registration mechanism. Given the small, known sample, duplicate entries were monitored manually, with 2 duplicates identified and the latest response considered the final one. |
| Handling of incomplete questionnaires | Were only completed questionnaires analyzed? Were questionnaires which terminated early (where, for example, users did not go through all questionnaire pages) also analyzed? | Mandatory response settings in Google Forms prevented incomplete submission; all submitted questionnaires were complete and included in analysis. |
| Questionnaires submitted with an atypical timestamp | Some investigators may measure the time people needed to fill in a questionnaire and exclude questionnaires that were submitted too soon. Specify the timeframe that was used as a cut-off point, and describe how this point was determined. | No timestamp-based exclusion criteria were applied. Survey 1 sessions were video-recorded, allowing verification that completion time was consistent with genuine engagement. Survey 2 was self-administered; no minimum completion time threshold was set, but given the small known sample and mandatory response enforcement, atypically rapid submissions were not identified as a concern. |
| Statistical correction | Indicate whether any methods such as weighting of items or propensity scores have been used to adjust for the non-representative sample; if so, please describe the methods. | No statistical correction for non-representative sampling (e.g., weighting, propensity scores) was applied, consistent with the exploratory and descriptive nature of the study. |
