## Supplementary File 3: COREQ checklist for "Leapfrogging cancer diagnostics with AI: a mixed-methods assessment of implementation determinants in low- and middle-income countries"

Consolidated criteria for reporting qualitative studies (COREQ): 32-item checklist

Developed from:

Tong A, Sainsbury P, Craig J. Consolidated criteria for reporting qualitative research (COREQ): a 32-item checklist for interviews and focus groups. International Journal for Quality in Health Care. 2007. Volume 19, Number 6: pp. 349 – 357

| **Item No** | | **Guide Questions/Description** | **Response** |  |  |
| --- | --- | --- | --- | --- | --- |
| **Domain 1: Research team and reflexivity** | | | |  |  |
| **Personal Characteristics** | | | |  |  |
| 1. Interviewer/ facilitator | | Which author/s conducted the interview or focus group? | Nanda Horeweg |  |  |
| 2. Credentials | | What were the researcher’s credentials? E.g., PhD, MD | PhD |  |  |
| 3. Occupation | | What was their occupation at the time of the study? | Clinical epidemiologist |  |  |
| 4. Gender | | Was the researcher male or female? | female |  |  |
| 5. Experience and training | | What experience or training did the researcher have? | Training in clinical epidemiology in Masters and PhD programs, including training in the design of quantitative and qualitative studies |  |  |
| **Relationship with participants** | | | |  |  |
| 6. Relationship established | | Was a relationship established prior to study commencement? | Two participants had prior academic collaboration with members of the study team, and one participant had met study team members at an academic conference; all other participants had no prior direct relationship with the investigators. |  |  |
| 7. Participant knowledge of the interviewer | | What did the participants know about the researcher? e.g. personal goals, reasons for doing the research? | Participants were informed of the study objectives and the research team's institutional affiliation via the recruitment email prior to participation. No participants had personal prior knowledge of the interviewer. Participants were not informed of the interviewer's specific personal motivations or theoretical assumptions beyond the stated study aims. |  |  |
| 8. Interviewer characteristics | | What characteristics were reported about the interviewer/facilitator? e.g. Bias, assumptions, reasons and interests in the research topic | The research team's interest in AI implementation in LMICs may have introduced implicit framing toward the topic's potential benefits; this was mitigated by the use of open-ended, non-leading questions and a semi-structured guide designed to elicit both facilitators and barriers. |  |  |
|  | **Domain 2: study design** | | |  |  |
|  | **Theoretical framework** | | |  |  |
|  | 9. Methodological orientation and Theory | What methodological orientation was stated to underpin the study? e.g. grounded theory, discourse analysis, ethnography, phenomenology, content analysis | Hybrid inductive-deductive thematic analysis, informed by the TICD framework for determinants of healthcare professional practice as an initial deductive scaffold, with codes and categories iteratively adapted from the data. |  |  |
|  | **Participant selection** | | |  |  |
|  | 10. Sampling | How were participants selected? e.g., purposive, convenience, consecutive, snowball | Purposive sampling via professional networks and targeted identification of clinicians and authors involved in cancer diagnostics and digital pathology research. |  |  |
|  | 11. Method of approach | How were participants approached? e.g., face-to-face, telephone, mail, email | Email invitation. |  |  |
|  | 12. Sample size | How many participants were in the study? | Nine participants across three FGD sessions (n=6, n=2, n=1); the single-participant session was analysed as an individual interview. |  |  |
|  | 13. Non-participation Setting | How many people refused to participate or dropped out? Reasons? | Of 15 individuals approached, 13 completed data collection. Three of the 12 analysed survey participants did not join an FGD due to scheduling constraints. No participants withdrew during FGD sessions. |  |  |
|  | 14. Setting of data collection | Where was the data collected? e.g., home, clinic, workplace | Online, via secure video-conferencing platform. |  |  |
|  | 15. Presence of nonparticipants | Was anyone else present besides the participants and researchers? | Yes, 1-3 non-participating members of the study team were present. |  |  |
|  | 16. Description of sample | What are the important characteristics of the sample? e.g. demographic data, date | Nine participants (6 pathologists, 2 gynecologist, 1 gynecologic oncologist) from nine countries across Africa, Asia, and Latin America; all affiliated with tertiary hospitals or private clinics. Full participant characteristics reported in Table 1. |  |  |
|  | **Data collection** | | |  | No |
|  | 17. Interview guide | Were questions, prompts, and guides provided by the authors? Was it pilot tested? | A semi-structured FGD guide was used, covering three primary questions with additional probes on regulatory, ethical, and implementation topics. The guide was developed by the research team and is provided as supplementary material. It was not formally pilot-tested prior to use. |  |  |
|  | 18. Repeat interviews | Were repeat interviews carried out? If yes, how many? | No repeat interviews were conducted. |  |  |
|  | 19. Audio/visual recording | Did the research use audio or visual recording to collect the data? | Sessions were audio and video recorded with participants' consent. |  |  |
|  | 20. Field notes | Were field notes made during and/or after the interview or focus group? | No formal field notes were taken during or after sessions. Transcripts served as the primary data record. |  |  |
|  | 21. Duration | What was the duration of the interviews or focus group? | 60-90 minutes per session. |  |  |
|  | 22. Data saturation | Was data saturation discussed? | Data saturation was not an aim of this exploratory pilot study; the sample was not designed or powered to achieve thematic saturation, and this is acknowledged as a limitation. |  |  |
|  | 23. Transcripts returned | Were transcripts returned to participants for comment and/or correction? | Transcripts were not returned to participants for review or correction. AI-assisted transcriptions were manually verified for accuracy by LAS. |  |  |
|  | **Domain 3: analysis and findings** | | |  |  |
|  | **Data analysis** | | |  |  |
|  | 24. Number of data coders | How many data coders coded the data? | Two coders (LAS, NH). |  |  |
|  | 25. Description of the coding tree | Did the authors provide a description of the coding tree? | An initial code tree was developed by LAS and refined jointly with NH prior to independent coding. The final code tree, comprising 39 codes across 7 domains, is provided in full as supplementary material. |  |  |
|  | 26. Derivation of themes | Were themes identified in advance or derived from the data? | Hybrid approach: an initial deductive structure informed by the TICD framework was applied, with codes and categories iteratively derived and adapted from the data. The final code tree reflects both pre-specified and inductively derived categories. |  |  |
|  | 27. Software | What software, if applicable, was used to manage the data? | QualCoder v3.7. |  |  |
|  | 28. Participant checking | Did participants provide feedback on the findings? | Yes, they reviewed the manuscript and provided feedback. |  |  |
|  | **Reporting** | | |  |  |
|  | 29. Quotations presented | Were participant quotations presented to illustrate the themes/findings? Was each quotation identified? e.g., participant number | Yes. Illustrative quotations are presented for major themes. Each quotation is identified by participant ID; country names and professions are not linked to individual quotations to preserve confidentiality given the small, geographically distinct sample. |  |  |
|  | 30. Data and findings consistent | Was there consistency between the data presented and the findings? | Yes, all themes reported in Results are grounded in coded transcript data; the full codebook and coded transcripts are available on request. |  |  |
|  | 31. Clarity of major themes | Were major themes clearly presented in the findings? | Major themes are reported under clearly labelled subheadings in Results, each with segment counts, participant counts, and illustrative quotations. |  |  |
|  | 32. Clarity of minor themes | Is there a description of diverse cases or a discussion of minor themes? | Minor and divergent themes were tracked separately during coding and are noted where relevant in Results. |  |  |
