## Supplementary File 4: Survey 1 instrument for "Leapfrogging cancer diagnostics with AI: a mixed-methods assessment of implementation determinants in low- and middle-income countries"

**Survey 1: Leapfrogging Cancer Diagnostics in LMICs with AI**

Goal Survey: Assess existing laboratory, imaging, pathology, oncology infrastructure for cancer diagnostics; map infrastructure, workflows, data readiness, AI opportunities, and barriers

### Basic information

1. **Country/ Region**: _____________
2. **Occupation:**
   - Pathologist
   - Oncologist
   - Specialist for laboratory medicine
   - Other (please specify) ____
3. **What is your sub-specialization or primary area of expertise:**

- Bone
- Breast
- Central Nervous System
- Dermatologic/Melanoma
- Endocrine/Thyroid
- Eye/Ocular
- Gastrointestinal
- Genitourinary
- Gynecologic
- Head & Neck
- Hematologic
- Hepatopancreatobiliary
- Neuro-oncology
- Oral & Maxillofacial
- Pediatric
- Sarcoma
- Skin (Non-melanoma)
- Soft Tissue
- Thoracic
- Other (specify): ______

4. **Years of Experience:**
   - 0-5
   - 6-10
   - 11-20
   - >20
5. **Position:**
   - Resident (Junior fellow)
   - Attending (Fellow)
   - Senior attending (Senior Fellow)
   - Lab director
   - Other: __________
6. **Current workplace(s)?**

Please provide:

- Institution name
- Your Position
- Type of Institution: Public hospital, Private hospital, Public lab or Private lab

### AI applications

We categorize AI applications as follows, and highlight applications in an early-stage *****, which require further research:

- **Screening & Early Detection:** e.g., Automated screening of PAP-smears (including cancer detection)
- **Cancer Detection & Diagnostic Models:** e.g., Cancer detection (e.g. in lymph nodes), Grading (Prostate Gleason Grading), Automated IHC Scoring (breast: Ki67, ER, PR, HER2, NSCLC: PD-L1), Mitosis detection and counting
- **Prediction of Biomarkers & Molecular Alterations from H&E:** e.g., MSI prediction for CRC, EGFR prediction in lung carcinoma
- **Risk Assessment (Relapse/ Prognosis):** e.g., relapse risk prediction in breast cancer, CRC prognosis prediction (favorable vs poor), prediction of distant recurrence risk in endometrial cancer
- ***** **Prediction of Treatment Response:** e.g. Neoadjuvant chemotherapy response prediction in NSCLC (non small cell lung cancer); Immunotherapy response prediction across cancer types
- ***** **Oncology "CoPilot”:** Combines pathology, radiology, clinical data to suggest next actions based on guidelines (e.g. which test to order, referral, treatment decisions)

1. **On a scale from 1 (Not important) to 5 (Extremely important), how important do you consider the following algorithm type for improving patient outcomes?**
   - Screening & Early Detection
   - Cancer Detection & Diagnosis
   - Treatment planning: Prediction of Biomarkers & Molecular Alterations from H&E
   - Cancer Treatment and follow-up: Risk Assessment (Relapse/ Prognosis)
   - * Cancer Treatment and follow-up: Prediction of Treatment Response
2. **When considering AI applications in pathology, there are often trade-offs between different factors. Imagine you have 100 points to allocate across the following five factors, according to how important each is for you when deciding on the adoption of an AI system. Please ensure your total adds up to 100.**
1. Performance (accuracy, reliability) → ___
2. Throughput (speed, scalability) → ___
3. Resource use (cost, staffing, infrastructure) → ___
4. Clinical utility (impact on patient outcomes, decision-making) → ___
5. Integration & usability (ease of workflow integration, user-friendliness) → ___
3. **For each AI application, what is the maximum level of automation you would currently feel confident adopting in clinical practice?**

0 = No role (AI not involved)

1 = Case triage (AI prioritizes or orders cases, pathologist still reviews all)

2 = Screening tool (AI rules out clearly negative/normal cases with high confidence, pathologist reviews positives and sample of negatives)

3 = Automatic predictions/decisions (AI provides a definitive diagnosis/prediction without pathologist review)

4 = AI agent (AI not only predicts/decides but also takes actions, e.g., orders additional tests, structures reports, orchestrates workflow)

**Applications:**

- Screening & Early Detection
- Cancer Detection & Diagnosis
- Prediction of Biomarkers & Molecular Alterations from H&E

4. **For each AI application, what is the maximum level of automation you would currently feel confident adopting in clinical practice?**

0 = No role (AI not involved)

1 = Automatic predictions/decisions (AI provides a definitive diagnosis/prediction without pathologist review)

2 = AI agent (AI not only predicts/decides but also takes actions, e.g., orders additional tests, structures reports, orchestrates workflow)

**Applications:**

- Risk Assessment (Relapse/Prognosis)
- * Prediction of Treatment Response

5. **If you could have only one application type, which one would it be and why?** _______________
6. **Are there other AI applications in pathology cancer diagnostics that you envision, which are not mentioned?** ___________________

### AI Requirements

1. **Rate your institution’s internet connectivity for digital pathology purposes** (e.g., uploading/downloading large slide files); 1 (Very Poor) – 5 (Excellent):
2. **Does your laboratory use a Laboratory Information System (LIS) for pathology data?**

☐ Yes, routinely ☐ Partially ☐ No

3. **Do you store pathology slides digitally (WSI or images archived in a Picture Archiving and Communication System (PACS))?**

☐ Yes ☐Partially, a selection of slides is stored ☐ No

4. **Please indicate which resources are available:**
   - Microscope
   - Phone
   - Computer
   - Microscope Camera
   - Low-Throughput WSI Scanner (<10 slides)
   - High-Throughput WSI Scanner (>10 slides)
   - Server (Storage, CPU compute)
   - Low-cost GPU (~$1500)
   - High performance cluster (Storage, GPUs)
   - Cloud (Rented remote storage)
   - Other: _____

If WSI Scanner is available, which vendor? ______________

Are there any resources that have not been captured but you would consider essential for AI integration?_________

### Feasibility of AI implementation

We provide three scenarios of how an AI Setup could practically look like:

| **Mobile** | Smartphone mounted on microscope using apps   - Pro’s: offline connectivity, highly accessible, low costs - Cons: poor image quality, no existing algorithms yet | 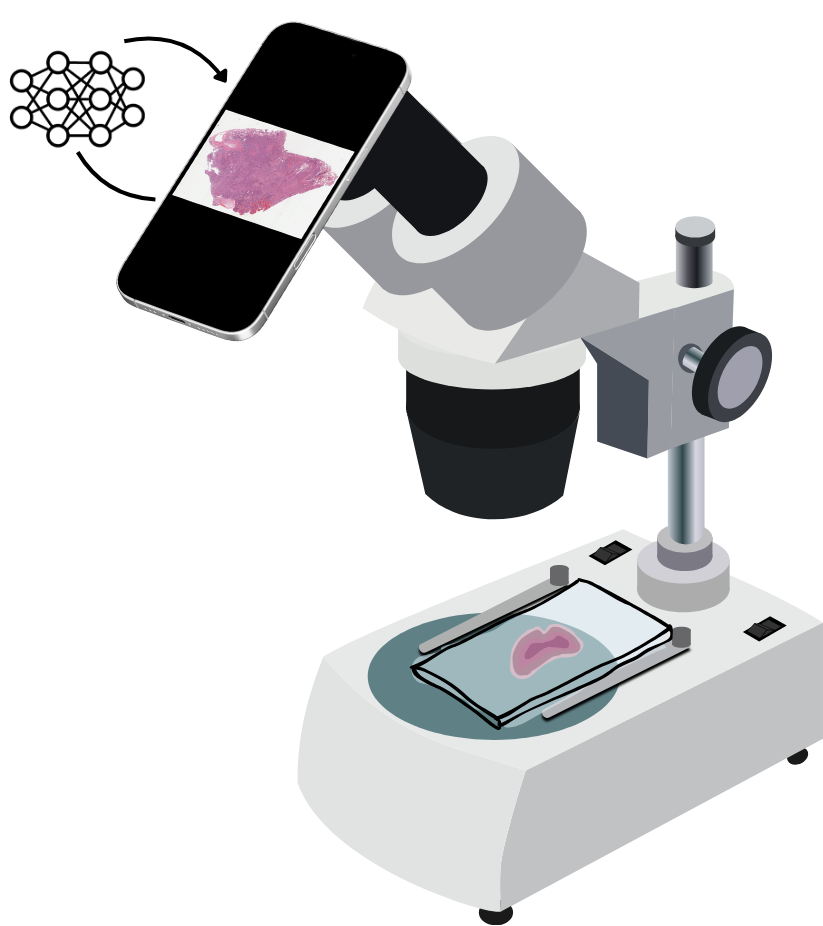 |
| --- | --- | --- |
| **Local workstation** | Microscope camera + WSI stitching software   - Pro’s: offline connectivity, more accessible than WSI scanners (~$3k for camera + software) - Cons: No existing algorithms, medium image quality | 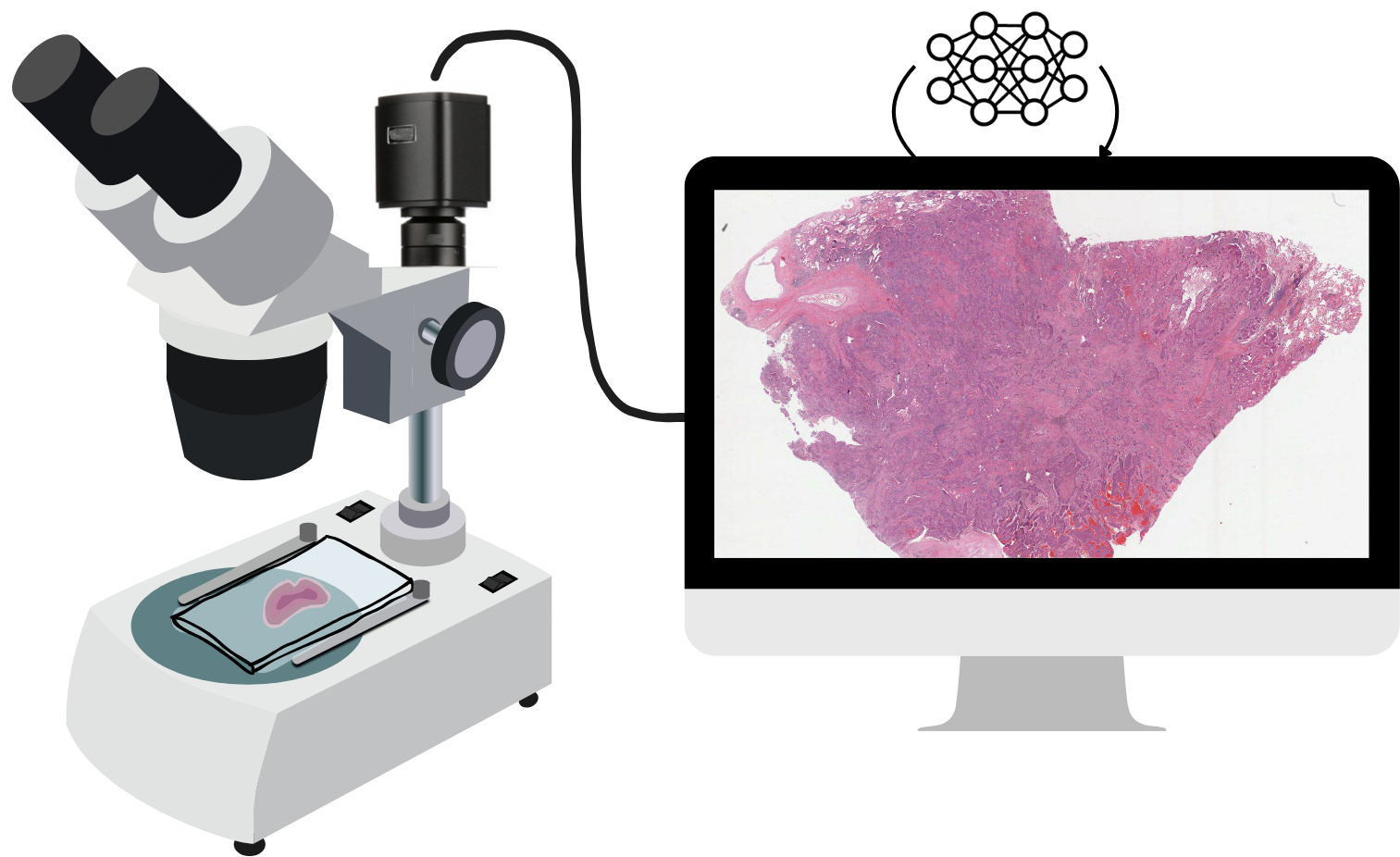 |
| **Cloud-based** | WSI scanner   - Pro’s: High image quality, existing cloud solutions and algorithms can run on remote cloud hardware - Cons: Internet required, low accessibility due to high costs from scanner | 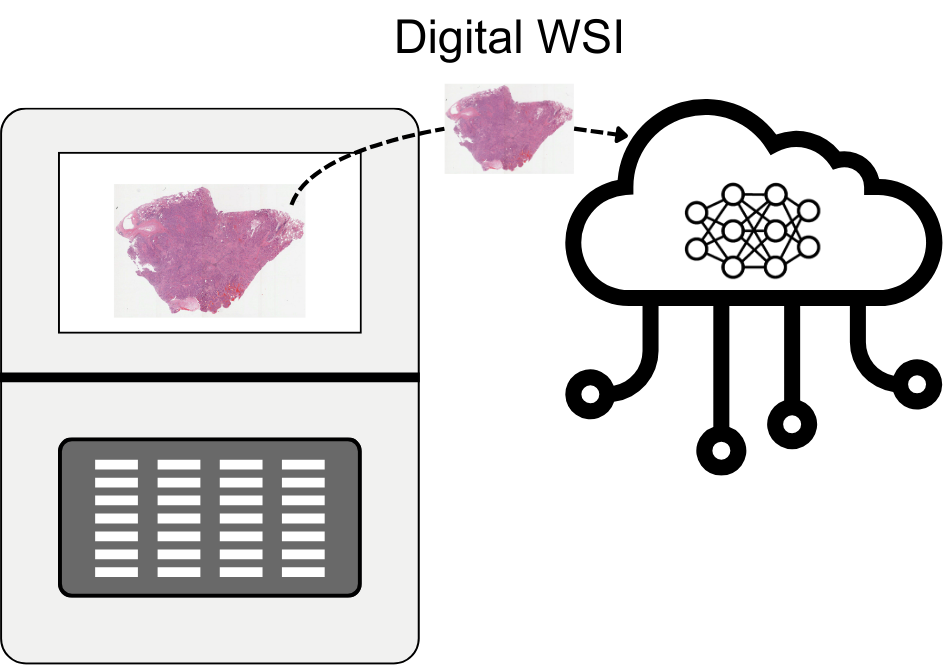 |

1. **Which setup would be the easiest to implement in your setting in the short term (≤1 year)?**

☐ Mobile

☐ Local workstation

☐ Cloud-based

☐ None

2. **For each scenario, how do you rate its feasibility (1: Not feasible, 5: Highly feasible) for implementation in your setting in the next 5-10 years?**

Mobile: ___ Local workstation: ____	Cloud: ___

3. **Do you envision other settings beyond the ones mentioned above?** ____________________
4. **If AI was implemented successfully, what would be the biggest improvement in your setting?**

☐ Faster turnaround time

☐ More accurate and standardized diagnoses

☐ Reduced workload for pathologists

☐ Cost savings (reduced need for physical slide delivery for remote consultation)

☐ Access to expert-level diagnostics in remote areas

☐ Other: __________

5. **What do you consider the three most critical barriers to AI adoption in your setting?**

- Workforce availability
- Personnel training / skills
- Financial constraints
- Hardware availability
- Connectivity limitations
- Availability of validated AI algorithms
- Data governance & security
- Interoperability (functionality of information systems to exchange data and enable sharing of information)
- Regulatory approval & reimbursement
- Other: _________

Which would be the single most critical barrier and why? ________

### Lab Infrastructure

1. **Are cytology-based cancer screening programs available in your country?**

☐ Yes ☐ No

2. **If yes, please specify which cancer screening programs: ________**
3. **Where are patient samples (cytology, histology) usually collected for testing?**

☐ Small health center or local clinic

☐ Mobile clinic or community program

☐ Large hospital or clinic

☐ Private lab

☐ Patients’ home

☐ Other (please specify) _________

4. **How long does it usually take from sample collection to fixation?**

☐ < 30min ☐ 30min -2 h ☐ 2-6h ☐ > 6 hours

5. **Which services/ tests are available?**

**Cytology**

☐ Pap smear ☐ FNA cytology ☐ Body fluids cytology ☐ Other: __________

**Histology**

**Is H&E staining routinely available?**

☐ Yes ☐ No

**If H&E staining is available, please rate its quality (1: Low, 5: High): ______**

**Is IHC staining available?**

- No IHC
- I have IHC, but it is extremely limited, hampering me to arrive at some WHO classification that requires IHC on a **daily** basis.
- I have IHC, but it is limited, hampering me to arrive at some WHO classification that requires IHC on a **weekly** basis.
- I have sufficient IHC panels to arrive at most WHO classifications that require IHC.

**Which number of IHC markers are available in your lab?**

- None
- 1-10
- 10-100
- >100

**Can you name a few IHCs that you feel are essential, but are missing in your lab?** ___________

**Are molecular tests available (select all that apply)?**

☐ Not available

☐ PCR / HPV DNA

☐ In-Situe Hybridisation (ISH)

☐ Hotspot techniques/ qPCR

☐ Targeted NGS (<100 genes)

☐ Advanced NGS (>100 genes)

☐ Other: ______

**Can you name a few molecular tests that you feel are essential, but are missing in your lab?** ___________

6. **Where are the following tests usually processed?**

**Cytology (Pap smears, FNAs):**
 ☐ In-house hospital lab ☐ Private lab ☐ Referral-only national lab
 ☐ Abroad (specify): ______ ☐ Not available

**Histology (biopsies, surgical specimens):**
 ☐ In-house hospital lab ☐ Private lab ☐ Referral-only national lab
 ☐ Abroad (specify): ______ ☐ Not available

**Immunohistochemistry:**
 ☐ In-house hospital lab ☐ Private lab ☐ Referral-only national lab
 ☐ Abroad (specify): ______ ☐ Not available

**Basic Molecular tests (i.e. PCR, ISH):**
 ☐ In-house hospital lab ☐ Private lab ☐ Referral-only national lab
 ☐ Abroad (specify): ______ ☐ Not available

**Advanced Molecular tests (i.e. NGS panel testing):**
 ☐ In-house hospital lab ☐ Private lab ☐ Referral-only national lab
 ☐ Abroad (specify): ______ ☐ Not available

7. **Turnaround Time: How long does it usually take to get results?**

**Cytology:** ☐ <1 day ☐ 1–3 days ☐ 4–7 days ☐ 1–2 weeks ☐ >2 weeks ☐ Not available

**Histology:** ☐ <1 day ☐ 1–3 days ☐ 4–7 days ☐ 1–2 weeks ☐ >2 weeks ☐ Not available

**Immunohistochemistry:** ☐ <1 day ☐ 1–3 days ☐ 4–7 days ☐ 1–2 weeks ☐ >2 weeks ☐ Not available

**Basic Molecular (i.e. PCR, ISH):** ☐ <1 day ☐ 1–3 days ☐ 4–7 days ☐ 1–2 weeks ☐ >2 weeks ☐ Not available

**Advanced Molecular (i.e. NGS panels):** ☐ <1 day ☐ 1–3 days ☐ 4–7 days ☐ 1–2 weeks ☐ >2 weeks ☐ Not available

8. **Pick the three most important factors limiting your laboratory’s quality:**

__ Tissue handling / fixation

__ Sectioning quality

__ Staining consistency

__ Equipment maintenance

__ Reagent/consumable stockouts

__ Reagent quality

__ Regulatory/procurement barriers

__ Lack of trained / specialized personnel

__ Data systems/LIS limitations
__ Space/infrastructure constraints

__ Other: __________________________

### Pathology Workforce availability

1. **Are pathologists available?**

- Yes, general and specialized pathologists
- Yes, general pathologists only
- Few general pathologists
- Not available

2. **On a scale from 1 (Very unsatisfactory ) to 5 (Very satisfactory) how satisfied are you with the number of available pathologists?**
3. **Please estimate the average workload per pathologist (number of cases per week)**

- <25 cases/week
- 25–50 cases/week
- 50–100 cases/week
- 100-500 cases/week
- >500 cases/week

4. **On a scale from 1 (Very unsatisfactory ) to 5 (Very satisfactory)**, **how satisfied are you with the workload?**
5. **OPTIONAL: For pathologists: Can you describe the biggest challenge you face in providing accurate cancer diagnoses? _____________**

### Oncology Treatment availability

1. **Rate the availability:**

**Trained oncologists:** ☐ Available routinely ☐ Available in selected centers ☐ Not available

**Trained oncological surgeons:** ☐ Available routinely ☐ Available in selected centers ☐ Not available

**Radiotherapy:** ☐ Available routinely ☐ Available in selected centers ☐ Not available

**Chemotherapy:** ☐ Available routinely ☐ Available in selected centers ☐ Not available

**Immunotherapy:** ☐ Available routinely ☐ Available in selected centers ☐ Not available

**Targeted therapies:** ☐ Available routinely ☐ Available in selected centers ☐ Not available

2. **If radiotherapy is available, which specifically?:**

☐ External beam radiotherapy ☐ Brachytherapy

3. **If targeted therapies are available, which specifically?**

- Anti-hormonal therapy
- HER2-targeted therapy
- EGFR/other targeted agents
- Don’t know
- Other: ________

**OPTIONAL: For oncologists: What is the biggest challenge you face in providing quality cancer care? _____________**

________________________________________________________________________________

**Final**

Thank you very much for your valuable insights and comments.

**Any other question(s) we should have asked?**

**Other comments?**
