## Supplementary File 5: Survey 1 guide for "Leapfrogging cancer diagnostics with AI: a mixed-methods assessment of implementation determinants in low- and middle-income countries"

### [TEMPLATE] Interview Conduct and Notes

**Interviewer:** LS / NvdB

**Interviewee:**

**Date&Time:**

#### 1. Introduction

Welcome and go through Introduction slides (until AI Applications) [Link Slides].

Make sure to ask if you can record and start the recording.

#### 2. Survey

Ask interviewee to start the survey [Link to survey] and share their screen.

| **Basic information** | **Notes** |
| --- | --- |
| Country/ Region: _____________ |  |
| Occupation:   - Pathologist - Oncologist - Specialist for laboratory medicine - Other (please specify) ____ |  |
| What is your sub-specialization or primary area of expertise:   - Bone - Breast - Central Nervous System - Dermatologic/Melanoma - Endocrine/Thyroid - Eye/Ocular - Gastrointestinal - Genitourinary - Gynecologic - Head & Neck - Hematologic - Hepatopancreatobiliary - Neuro-oncology - Oral & Maxillofacial - Pediatric - Sarcoma - Skin (Non-melanoma) - Soft Tissue - Thoracic - Other (specify): ______ |  |
| Years of Experience:   - 0-5 - 6-10 - 11-20 - >20 |  |
| Position   - Resident (Junior fellow) - Attending (Fellow) - Senior attending (Senior Fellow) - Lab director - Other: __________ |  |
| Current workplace(s)?  *[Conduct Note]: Verify they enter all required information, as this is free-text entry. Ensure they add all current workplaces.*  Please add:   - Institution name - Your Position - Type of Institution: Public hospital, Private hospital, Public lab or Private lab |  |

Before continuing the AI Applications section introduce the different types of applications with the slides [Link to Slides].

| **AI Applications** | **Notes** |
| --- | --- |
| On a scale from 1 (Not important) to 5 (Extremely important), how important do you consider the following algorithm type for improving patient outcomes?  *[Conduct Note]: Explain that the * highlights applications that are at an early stage of development (No CE-IVD certified examples, no existing clinical implementation.)*   - Screening & Early Detection - Cancer Detection & Diagnosis - Treatment planning: Prediction of Biomarkers & Molecular Alterations from H&E - Cancer Treatment and follow-up: Risk Assessment (Relapse/ Prognosis) - * Cancer Treatment and follow-up: Prediction of Treatment Response |  |
| When considering AI applications in pathology, there are often trade-offs between different factors. Imagine you have 100 points to allocate across the following five factors, according to how important each is for you when deciding on the adoption of an AI system. **Please ensure your total adds up to 100.**  *[Conduct Note]: Verify they enter correct numbers as this is free-text entry.*   - Performance (accuracy, reliability) - Throughput (speed, scalability) - Resource use (cost, staffing, infrastructure) - Clinical utility (impact on patient outcomes, decision-making) - Integration & usability (ease of workflow integration, user-friendliness) |  |
| For each AI application, what is the maximum level of automation you would currently feel confident adopting in clinical practice?  *[Conduct Note]: Explain the different roles:*  0 = No role (AI not involved)  1 = Case triage (AI prioritizes or orders cases, pathologist still reviews all)  2 = Screening tool (AI rules out clearly negative/normal cases with high confidence, pathologist reviews positives and sample of negatives)  3 = Automatic predictions/decisions (AI provides a definitive diagnosis/prediction without pathologist review)  4 = AI agent (AI not only predicts/decides but also takes actions, e.g., orders additional tests, structures reports, orchestrates workflow)  *[Conduct Note]: Only applications which replicate what pathologists do or provide predictions on an existing biomarker.*  Applications:   - Screening & Early Detection - Cancer Detection & Diagnosis - Prediction of Biomarkers & Molecular Alterations from H&E |  |
| For each AI application, what is the maximum level of automation you would currently feel confident adopting in clinical practice?  0 = No role (AI not involved)  1 = Automatic predictions/decisions (AI provides a definitive diagnosis/prediction without pathologist review)  2 = AI agent (AI not only predicts/decides but also takes actions, e.g., orders additional tests, structures reports, orchestrates workflow)  *[Conduct Note]: Only applications which provide a novel biomarker.*  Applications:   - Risk Assessment (Relapse/Prognosis) - * Prediction of Treatment Response |  |
| If you could have only one application type, which one would it be and why? ___________ |  |
| Are there other AI applications in pathology cancer diagnostics that you envision, which are not mentioned? ___________________ |  |

| **AI Requirements** | **Notes** |
| --- | --- |
| Rate your institution’s internet connectivity for digital pathology purposes (e.g., uploading/downloading large slide files); 1 (Very Poor) – 5 (Excellent) |  |
| Does your laboratory use a Laboratory Information System (LIS) for pathology data?  ☐ Yes, routinely ☐ Partially ☐ No |  |
| Do you store pathology slides digitally (WSI or images archived in a Picture Archiving and Communication System (PACS))?  *[Conduct Note]:* A Picture Archiving and Communication System (PACS) is a digital medical imaging technology that acquires, stores, distributes, and displays medical images from various modalities like whole slide images, X-rays, CT scans, and MRIs.  ☐ Yes ☐Partially, a selection of slides is stored ☐ No |  |
| Please indicate which resources are available:   - Microscope - Phone - Computer - Microscope Camera - Low-Throughput WSI Scanner (<10 slides) - High-Throughput WSI Scanner (>10 slides) - Server (Storage, CPU compute) - Low-cost GPU (~$1500) - High performance cluster (Storage, GPUs) - Cloud (Rented remote storage) - Other: _____ |  |
| If WSI Scanner is available, which Vendor? |  |
| Are there any resources that have not been captured but you would consider essential for AI integration?_________ |  |

**Feasibility of AI implementation**

Go through three scenarios of how an AI Setup could practically look like:

| **Mobile** | Smartphone mounted on microscope using apps   - Pro’s: offline connectivity, highly accessible, low costs - Cons: poor image quality, no existing algorithms yet | 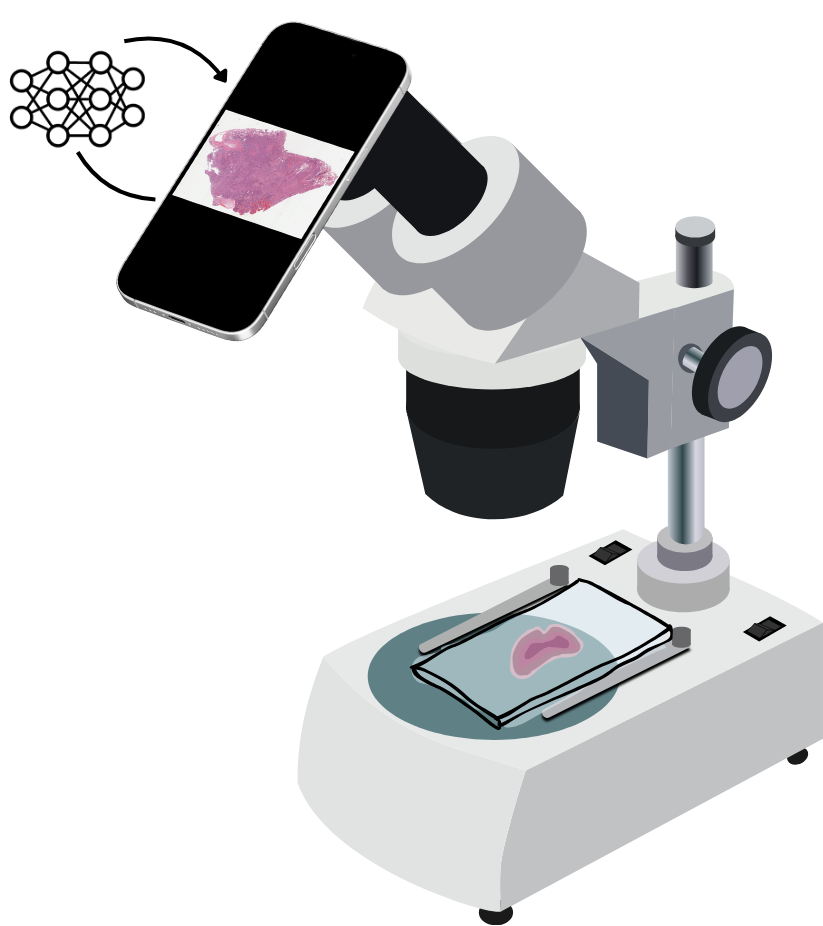 |
| --- | --- | --- |
| **Local workstation** | Microscope camera + WSI stitching software   - Pro’s: offline connectivity, more accessible than WSI scanners (~$3k for camera + software) - Cons: No existing algorithms, medium image quality | 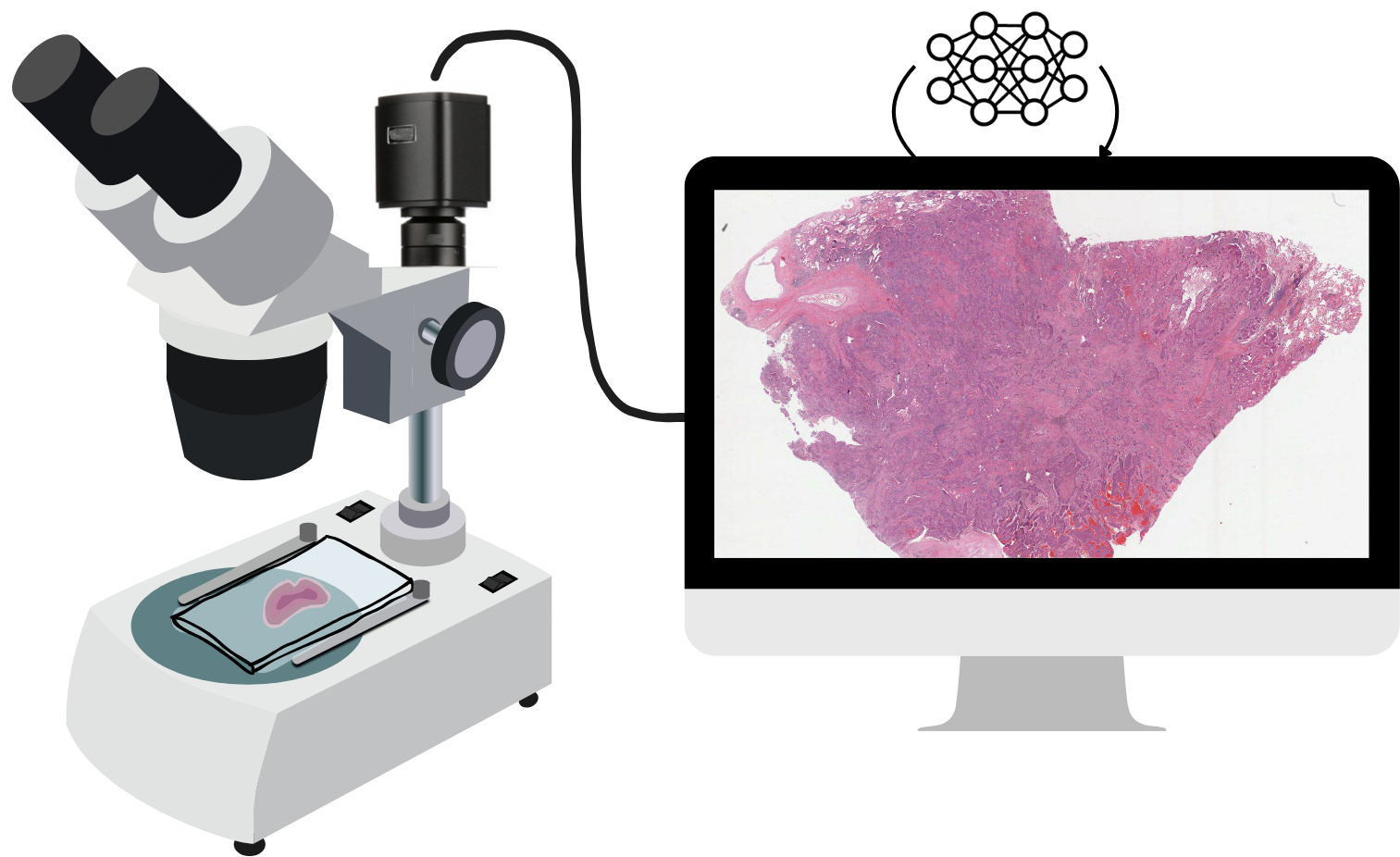 |
| **Cloud-based** | WSI scanner   - Pro’s: High image quality, existing cloud solutions and algorithms can run on remote cloud hardware - Cons: Internet required, low accessibility due to high costs from scanner | 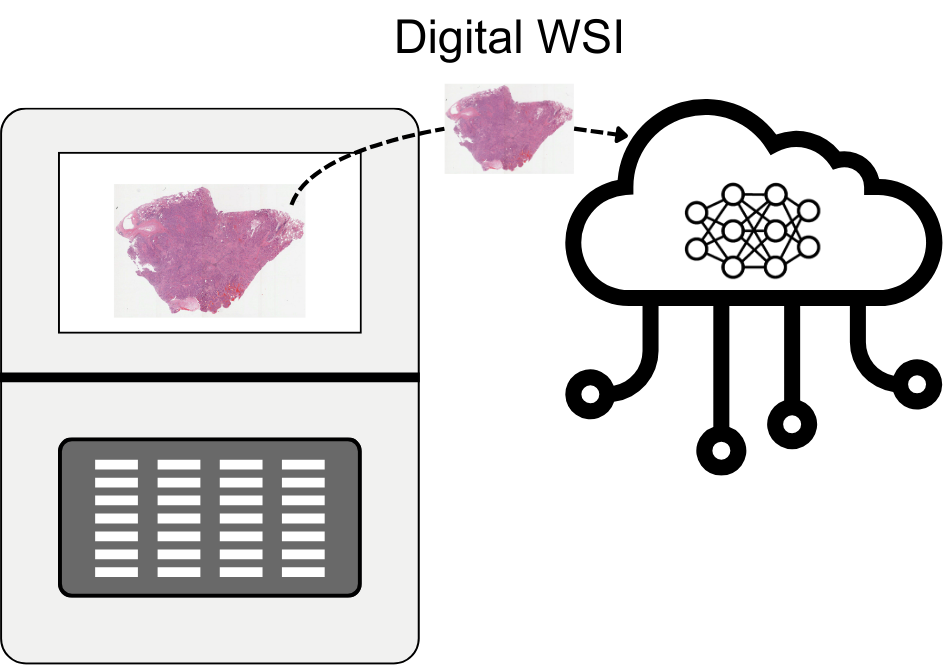 |

| **Feasibility of AI implementation** | **Notes** |
| --- | --- |
| Which setup would be the easiest to implement in your setting in the short term (≤1 year)?  ☐ Mobile  ☐ Local workstation  ☐ Cloud-based  ☐ None |  |
| For each scenario, how do you rate its feasibility (1: Not feasible, 5: Highly feasible) for implementation in your setting in the next 5-10 years?  Mobile: ___ Local workstation: ____ Cloud: ___ |  |
| Do you envision other settings beyond the ones mentioned above? ____________________ |  |
| If AI was implemented successfully, what would be the biggest improvement in your setting?  ☐ Faster turnaround time  ☐ More accurate and standardized diagnoses  ☐ Reduced workload for pathologists  ☐ Cost savings (reduced need for physical slide delivery for remote consultation)  ☐ Access to expert-level diagnostics in remote areas  ☐ Other: __________ |  |
| What do you consider the three most critical barriers to AI adoption in your setting?  *[Conduct Note]: Ensure they only tick 3 boxes.*   - Workforce availability - Personnel training / skills - Financial constraints - Hardware availability - Connectivity limitations - Availability of validated AI algorithms - Data governance & security - Interoperability (functionality of information systems to exchange data and enable sharing of information) - Regulatory approval & reimbursement - Other: _________ |  |
| Which would be the single most critical barrier and why? ________ |  |

| **Lab Infrastructure** | **Notes** |
| --- | --- |
| Are cytology-based cancer screening programs available in your country?  ☐ Yes ☐ No |  |
| If yes, please specify which cancer screening programs: ____________ |  |
| Where are patient samples (cytology, histology) usually collected for testing?  ☐ Small health center or local clinic  ☐ Mobile clinic or community program  ☐ Large hospital or clinic  ☐ Private lab  ☐ Patients’ home  ☐ Other (please specify) _________ |  |
| How long does it usually take from sample collection to fixation?  ☐ < 30min ☐ 30min -2 h ☐ 2-6h ☐ > 6 hours |  |
| Which cytology services/ tests are available?  ☐ Pap smear ☐ FNA cytology ☐ Body fluids cytology ☐ Other: __________ |  |
| Is H&E Staining routinely available?  ☐ Yes ___ ☐ No |  |
| If H&E staining is available, please rate its quality (Scale 1: Low - 5: High): ____ |  |
| Is IHC staining available?   - No IHC - I have IHC, but it is extremely limited, hampering me to arrive at some WHO classification that requires IHC on a **daily** basis. - I have IHC, but it is limited, hampering me to arrive at some WHO classification that requires IHC on a **weekly** basis. - I have sufficient IHC panels to arrive at most WHO classifications that require IHC. |  |
| Which number of IHC markers are available in your lab?   - None - 1-10 - 10-100 - >100 |  |
| Can you name a few IHCs that you feel are essential, but are missing in your lab? ___________ |  |
| Are molecular tests available (select all that apply)?  ☐ Not available  ☐ PCR / HPV DNA  ☐ In-Situe Hybridisation (ISH)  ☐ Hotspot techniques/ qPCR  ☐ Targeted NGS (<100 genes)  ☐ Advanced NGS (>100 genes)  ☐ Other: ______ |  |
| Can you name a few molecular tests that you feel are essential, but are missing in your lab? ___________ |  |
| Where are the following tests usually processed?  *[Conduct Note]: If available abroad, note down which country/countries.*  In-house hospital lab ☐ Private lab  ☐ Referral-only national lab ☐ Abroad ☐ Not available  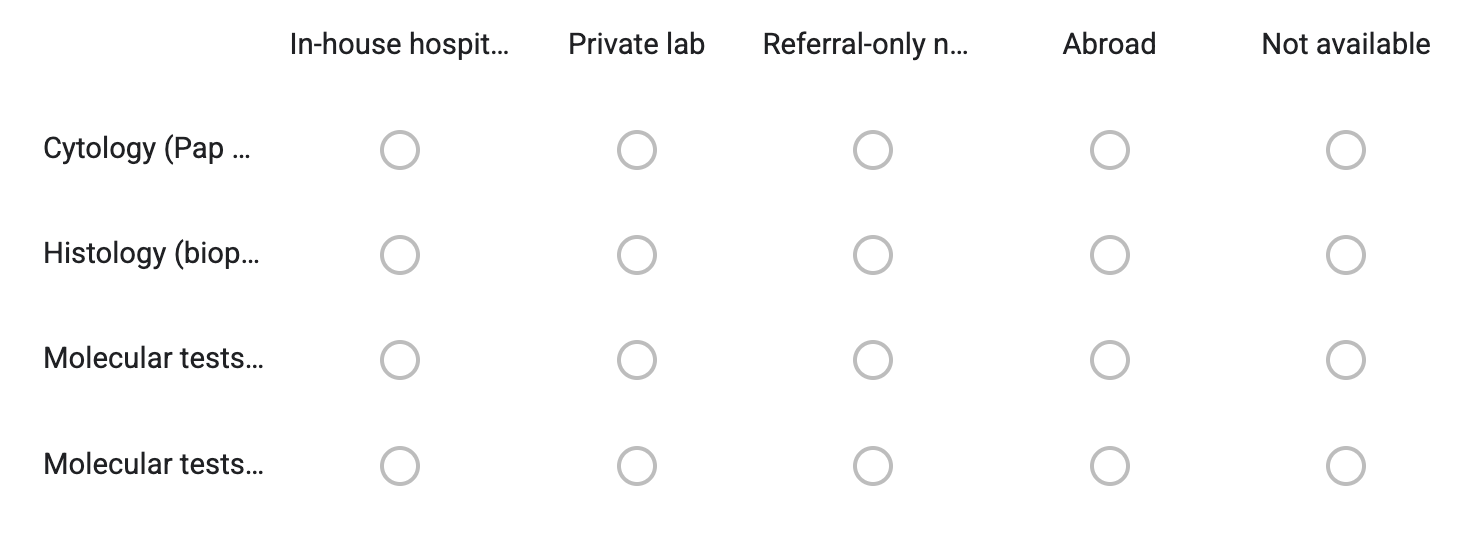 |  |
| Turnaround Time: How long does it usually take to get results?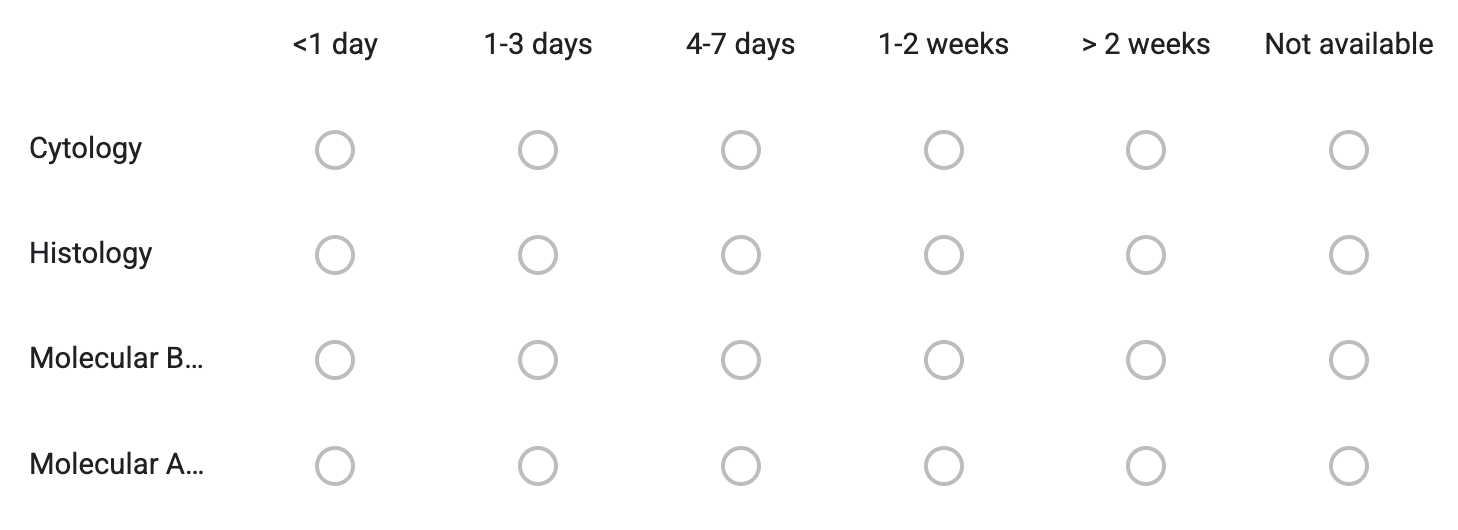 |  |
| Pick the three most important factors limiting your laboratory’s quality:  *[Conduct Note]: Ensure they only tick 3 boxes.*  __ Tissue handling / fixation  __ Sectioning quality  __ Staining consistency  __ Equipment maintenance  __ Reagent/consumable stockouts  __ Reagent quality  __ Regulatory/procurement barriers  __ Lack of trained / specialized personnel  __ Data systems/LIS limitations  __ Space/infrastructure constraints  __ Other: __________________________ |  |

| **Pathology Workforce availability** | **Notes** |
| --- | --- |
| Are pathologists available?   - Yes, general and specialized pathologists - Yes, general pathologists only - Few general pathologists - Not available |  |
| On a scale from 1 (Very unsatisfactory ) to 5 (Very satisfactory) how satisfied are you with the number of available pathologists? |  |
| Please estimate the average workload per pathologist (number of cases per week)   - <25 cases/week - 25–50 cases/week - 50–100 cases/week - 100-500 cases/week - >500 cases/week |  |
| On a scale from 1 (Very unsatisfactory ) to 5 (Very satisfactory), how satisfied are you with the workload? |  |
| **OPTIONAL:** For pathologists: Can you describe the biggest challenge you face in providing accurate cancer diagnoses? _____________  *[Conduct Note]: Only discuss if you have time left.* |  |

| **Oncology Treatment availability** | **Notes** |
| --- | --- |
| Rate the availability: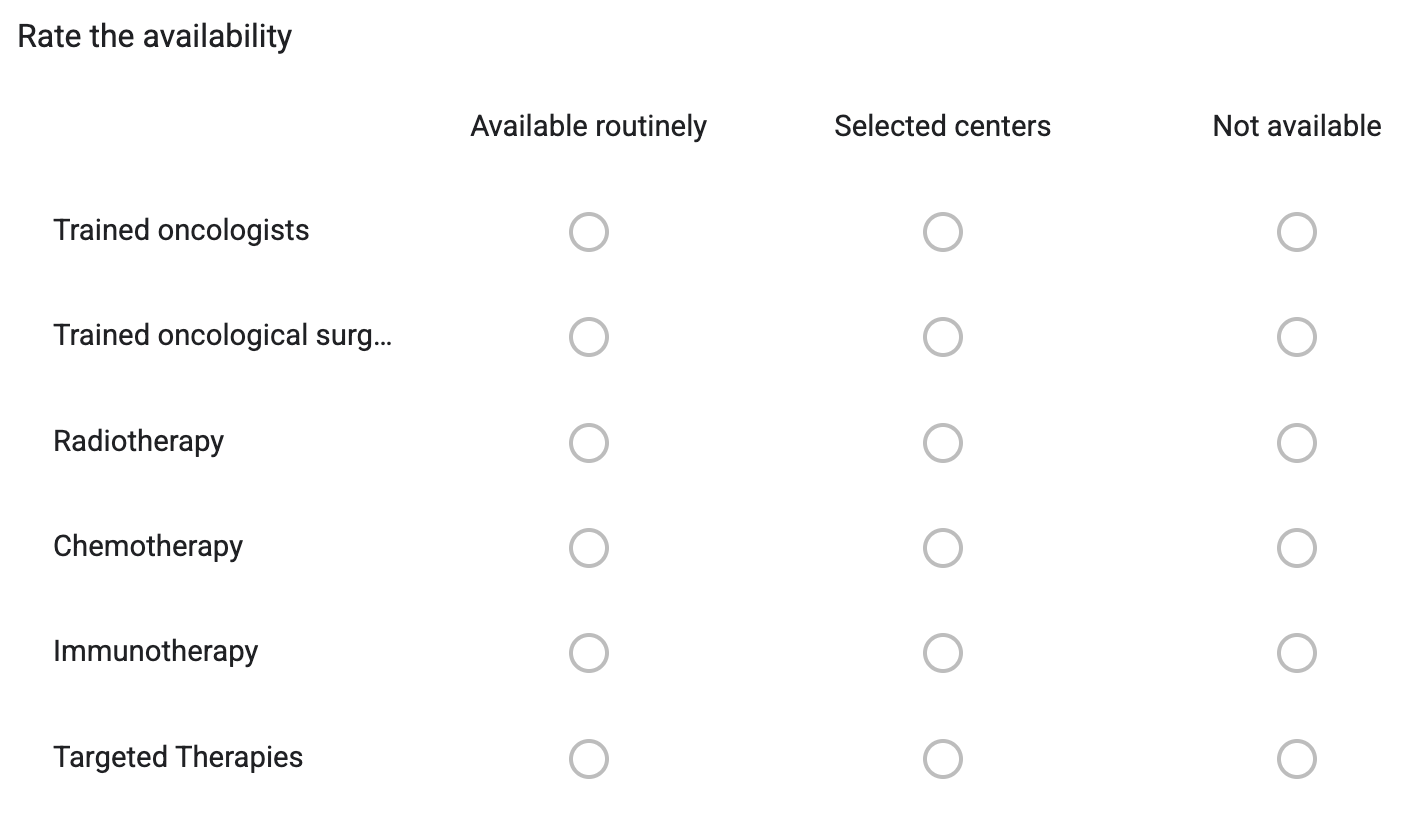 |  |
| If radiotherapy is available, which specifically?  ☐ External beam radiotherapy ☐ Brachytherapy |  |
| If targeted therapies are available, which specifically?   - Anti-hormonal therapy - HER2-targeted therapy - EGFR/other targeted agents - Other: ________ - Don’t know |  |
| OPTIONAL: For oncologists: What is the biggest challenge you face in providing quality cancer care? _____________  *[Conduct Note]: Only discuss with oncologists if you have time left.* |  |

End of official survey part, can share feedback now, or provide feedback afterwards e.g. via mail. **Ensure they submit the survey.**

*[Conduct Note]: OPTIONAL, no need to go through, only if desired by interviewee.*

| **Feedback questions** | **Notes** |
| --- | --- |
| Any other question(s) we should have asked? |  |
| Other comments? |  |

#### 3. Official end of survey

Stop recording, thank them for their time and inputs. Go through “What’s next?” slides [Link to Slides].

Which date would they prefer?
