## Supplementary File 7: Survey 2 instrument for "Leapfrogging cancer diagnostics with AI: a mixed-methods assessment of implementation determinants in low- and middle-income countries"

**AI Awareness & Attitude**

**1. How would you rate the average current level of awareness and use of AI in your country?**

- Absent	(not aware)
- Early	(aware of AI, but not using AI)
- Moderate	(incidental use of AI)
- Well underway	(common use of AI)
- Fully established	(use of AI fully integrated)

**2. How would you rate the average attitude or perception of healthcare providers toward use of AI in pathology in your country?**

- Very negative	(actively opposing AI implementation)
- Negative	(negative opinion or suspiciousness, but no action)
- Neutral	(neutral opinion and no action in favor or against)
- Positive	(positive opinion or curiosity, but no action)
- Very positive	(actively promoting AI implementation)

**3. How would you rate the average attitude or perception of cancer patients toward use of AI as part of their clinical journey in your country?**

- Very negative	(actively opposing AI usage)
- Negative	(negative opinion or suspiciousness, but no objection)
- Neutral	(neutral opinion, neither in favor nor against AI usage)
- Positive	(positive opinion or curiosity, but not demanding it)
- Very positive	(actively demanding AI usage)

**AI Literacy & Education**

**4. To what extent is education on AI currently integrated in the primary, secondary and higher education systems of your country?**

- Absent	(not at all)
- Early	(awareness of the need, but nothing realized yet)
- Moderate	(incidental, non-structural education on AI present)
- Well underway	(formally integrated at some levels of education)
- Fully established	(fully integrated across all levels of education)

**5. To what extent is education on AI for medical applications currently provided to health care providers in your country?**

- Absent	(not at all)
- Early	(awareness of the need, but nothing realized yet)
- Moderate	(incidental, non-structural education on AI present)
- Well underway	(formally integrated at some locations)
- Fully established	(fully integrated across the country)

**6. On which topics would additional education for health care providers in your country be most valuable? (Select max 3)**

- Basic AI concepts
- Digital pathology workflows
- AI implementation in clinical practice
- AI ethics and governance
- Regulatory approval pathways
- Interpretation of AI research papers
- Grant writing / research collaboration
- Technical development and programming
- Data management and annotation
- Validation of AI algorithms

**7. Which educational formats would be most feasible for health care providers in your country? (Select max 3)**

- Integration into clinical (pathology) residency
- Short in-person courses
- Conference workshops
- Regional training hubs
- Self-paced online modules
- Live online workshops
- International fellowships
- Mentorship programs

**AI Governance, Ethics and Funding**

**8. How would you rate your country’s current readiness regarding AI governance and regulation?**

- Absent	(no laws/regulations available yet)
- Early	(being developed now, expected within a year)
- Moderate	(general laws/regulations available, not specific for health care)
- Well underway	(laws/regulations specifically for health care available)
- Fully established	(laws/regulations specifically for health care available and regularly updated)

**9. How would you rate your country’s current readiness regarding AI ethics?**

- Absent	(no guidelines available yet)
- Early	(being developed now, expected within a year)
- Moderate	(general guidelines available, not specific for health care)
- Well underway	(guidelines specifically for health care available)
- Fully established	(guidelines specifically for health care available and regularly

updated)

**10. Which funding sources are currently most realistic for AI implementation in your country? (Select max 3 if any)**

- Government and public funding
- Insurance/reimbursement models
- Private healthcare investment
- Industry partnerships
- Academic research funding
- International grants
- Philanthropic/global health funding

**Stakeholders & Collaboration**

**11. Rank the 5 most important stakeholders for AI implementation in your country (Rank max 5)**

- Government
- Public hospitals/labs
- Private hospitals/labs
- Clinicians and other health care providers
- Professional medical societies
- (Academic) research groups
- International research partners
- Industry / AI companies
- Global health organizations
- Patients and patient organizations
- General Public

**12.** Which collaboration would you consider **most important** to advance AI in pathology in your country? Please list three types of collaborations and name **both stakeholders and type**.

**13.** Which collaborations are needed but **currently missing** to advance AI in pathology in your country? Please list three types of collaborations and name **both stakeholders and type**.

**14. What is the biggest barrier to sustaining collaboration in your country? (Select one)**

- Fragmented institutions / no existing networks
- Regulatory or legal obstacles (e.g. data governance, sample export restrictions)
- Dependence on short-term grants or donor cycles
- No shared data standards or infrastructure
- Competing institutional priorities
- Other: ___

**Clinical implementation**

**15. AI and digital pathology could change where each step of the diagnostic workflow takes place. In a hub-and-spoke model, where do you envision each step happening in your country?**

*(Spoke = district/rural hospital; Hub = regional or national referral center)*

|  | **Spoke *(close to patient)*** | **Hub *(referral center)*** |
| --- | --- | --- |
| **Sample preparation** (cutting, staining) | ☐ | ☐ |
| **Digitization** (slide scanning) | ☐ | ☐ |
| **AI analysis** | ☐ | ☐ |
| **Pathologist review & reporting** | ☐ | ☐ |

**16. To what extent do you think local studies *(e.g. validating AI performance on local patient data before clinical use)* are required before implementation of AI models in healthcare in your country?**

- Not required	(implementation of foreign AI models is sufficient)
- Desired, but not required	(preferrable if possible, but not a hard requirement)
- Required	(only locally validated models should be implemented)

**17. If you could prioritize ONE action to accelerate responsible AI implementation**

**in pathology in your country, what would it be?**

(open text)
