## Supplementary File 9: Focus group discussion guide for "Leapfrogging cancer diagnostics with AI: a mixed-methods assessment of implementation determinants in low- and middle-income countries"

Supplementary File 9: Focus Group Discussions Guide

### Introduction - TB

**XX:30-XX:33:** Wait for everyone to arrive, **start recording @LAS**

**XX:33-XX:35: Welcome,** info that we are recording the sessions, Thank everyone for participating in the surveys, (briefly) bring the purpose back to everyone's mind

**XX:35-XX:38:** Introduce everyone by name, profession and country, ask them to wave when mentioned

**XX:38-XX:39**: Briefly go over agenda:

- Questions and Feedback (~7min)
- Moderated discussion round (~40min)
- What’s Next? (~5min)

**XX:39-XX:40:** Handover to LAS

### Survey 1 results + Q&A - LAS

**XX:40-XX:50:** Overview on survey 1 participants, desired AI applications and settings, opportunities and barriers, current infrastructure results

**XX:50-XX:51:** Invite questions and feedback

**XX:51-XX:58:** Q&A

**XX:58:** End Q&A, handover to NH as moderator of focus group discussion

### Focus group discussion - NH

**XX:58-XX:00:** Introduce moderated discussion as part of data collection for paper

- **Goal:** Understand “what would it take to introduce AI into cancer diagnostics in LMICs?”
- There are no “wrong answers”: Feel free to comment, even if your thoughts, ideas and experiences contradict those of others
- We might need to cut someone off if it gets too long/ imbalanced OR ask quieter participants to add their viewpoint, and we will lead through the different questions, but are not joining the discussion itself - goal is for participants to discuss with each other

Keep track of who has contributed, encourage equal participation where possible:

|  | **Contributed to** | | |
| --- | --- | --- | --- |
| **Focus group participants DD.MM.YYYY** | **Question 1** | **Question 2** | **Question 3** |
| P01 |  |  |  |
| P02 |  |  |  |
| P03 |  |  |  |
| … |  |  |  |
| PXX |  |  |  |

**XX:00-XX:25:** Focus Group

Prompts:

1. Imagine we start an AI project in cancer diagnostics tomorrow. What are the first steps that need to happen?

Follow-up: Which steps are easiest vs most challenging?; Which steps require local vs external support?

2. Who are the key actors and facilitators needed, and what should they do?

Follow-up: Who needs to be involved right at the start, and who becomes important later on?; Which actors/ institutions might hinder progress?

3. What types of collaboration do you envision?

Follow-up: What would good collaboration look like in practice (compared to “in principle”)?; How could you collaborate with each other, but also with actors from HIC?

**XX:23-XX:25:** Wrap up, thank them for participating, handover to LAS for “What’s Next?”

### What’s Next? - LAS

**XX:25-XX:30:** Next steps for this project, but also clearly state our openness for future collaboration, thank them again on behalf of the whole team, looking forward to their feedback on the paper
