## Supplementary File 9: Focus group discussion code book for "Leapfrogging cancer diagnostics with AI: a mixed-methods assessment of implementation determinants in low- and middle-income countries"

Supplementary File 10: Focus group discussion codebook

Category: AI literacy

This category captures the social and educational preconditions for AI adoption, the enabling mindset of professionals, patients, and the public to meaningfully engage with AI tools.

...█Code: Attitudes and acceptability

Individual or group opinions, trust, concerns, or evaluations about AI, including its adoption, usefulness, safety, or ethical implications.

- “ I'm glad to say that in Malaysia the training of pathologists and scientists mindset towards AI is now pretty good”
- “I would assume that there is a general positivity into adoption of AI”
- “And also, around pathologists the perception around AI and cancer diagnostics needs to be palpated and felt. What does it mean to the pathologists? How are they going to view this? Is it a panacea? Is it a magic bullet that is coming in? Or is it a tool that is going to help? To solve the issues we are already grappling with”

...█Code: Awareness and sensitisation:

Population- or group-level knowledge of AI's existence and relevance

- “Now if these players come in we have to also begin to sensitise the public about the potentials of AI”
- “So, therefore that raises issues of literacy so we have to go back and re-educate as university people educate the people on the real potential, solve the myths around AI”
- “So what to me, I think is important is to really make all people aware, contribute positively, because now it's kind of segmented.”

...█Code: Educational integration

Formal or structured inclusion of AI knowledge, tools, or skills within educational curricula or training programs. NOT directly related to training healthcare professionals as part of an AI implementation.

- “now AI is a compulsory course for all university students”
- “the knowledge and information on AI should therefore be inculcated in the curricula of the various educational levels in Uganda”
- “we are pushing for more masters and PhDs in AI.”

Category: Evidence generation

This determinant captures the perceived need for research demonstrating AI's clinical benefit.

...█Code: Research and pilot studies

Mentions of broader applied research (research projects/ pilot studies) as establishing feasibility and clinical utility of AI applications, or the need for such projects.

- “You know, like the main leg, the principal actor here, it's university hospitals, as they can usually start producing data to bring up the whole different, for example, in this case, pathologists to adopt one new technology. And also because in the university hospitals, you can produce the research that everyone is talking about to give numbers, to make this something feasible”
- “we have the people trying to train AI models and even some research that are going on.”

...█Code: Clinical AI validation

Comments regarding validation of AI tools' performance in local populations and workflows before clinical use.

- “And a very important aspect I want to comment on validation of the results that is channeled by AI, because if these results are not validated then we should pick up anything that is at variance of the standard in terms of diagnosis then people might lose interest in this AI-assisted cancer diagnosis and most people may not even patronize it at all.”

Category: Governance frameworks

This category captures perceived needs and gaps in policy, regulation, and ethical frameworks governing AI use in clinical practice.

...█Code: Development of guidelines

Mentions of developing professional/clinical guidelines for acceptable AI use.

- “The second was to develop a consensus on guidelines and regulations as to what is acceptable in the histopathology report. How much of AI assisted work can be accepted within the report.”

...█Code: Development of policies/ legislation

Mentions of government-issued strategy and law regarding AI usage.

- “because it is on the side of government that we shall get the right legislation coming into parliament and empowering the laws that will govern AI.”
- “the government released an AI strategy”

...█Code: Ethics/ Consent, Count: 16

Comments discussing ethical questions such as patient safety, data privacy and consent.

- “When it comes to consent matters, we integrate consent for use of this service together with clinical consent. So when the patient makes consent for surgery, they consent to have pathology done. And this may include the very early AI that we're using right now.”
- “people ask questions. So what happens to the data? How safe is our data? And things like that.”

...█Code: Regulatory readiness / gaps

Mentions of the adequacy and pace of existing regulatory frameworks.

- “the policies and the regulatory issues in as far as AI in diagnostics are not yet ready. From my further reading recently, I have noted that the AI laws are expected, I think, in this year, 2026.”
- “who is liable for this? There are no laws as yet.”

Category: Clinical deployment

This determinant captures perceived requirements for deploying AI tools in clinical routine.

...█Code: Infrastructure:

Comments related to existing or required infrastructure, can be digitization (WSI scanners etc), clinical (pathology related), internet, road, technical (AI equipment).

- “As we say garbage in garbage out, so if we have a good gynecologist who can bring in good sample then we have a pathologist or scientist who can really make a good staining of the tissue”
- “Yeah, the internet connectivity is good. It's pretty good throughout the country”
- “small scanner whereby they can send images it is getting interpreted here.”

...█Code: Deployment strategy:

Deployment strategies for AI, e.g. how to integrate into existing pathways/ structures.

- “if there was a way to, say, distribute scanners, there would be labs that would be able to get to that point where they can scan images and then now they can be shared to the few available, um, let's say it's a pathologist who's going to report, that can be done.”
- “And therefore also empowerment of the practitioner. But whereas now the brains are congregated in one or two places. We would still want to see people going out and practicing at the different levels. And only be able to consult centrally on difficult cases. But offer largely a good service on easy cases.”

...█Code: Equity of access and current healthcare landscape

Comments related to inequity/ challenges/ imbalances in patients accessing the healthcare systems; can be geographical or financial hurdles, as well as assessment of current geographical distribution/ setup of cancer diagnostics/ pathology

- “So it's possible to, to bridge those distance gaps or human resource gaps using tech. And I think AI would come in there”
- “And most of the diagnostics, if you want quality, you want to run to the private sector.”
- “So then we can have a fair distribution of the service.”

...█Code: Pathologist Workforce:

Comments on current pathologist workforce availability and potential future risks/ opportunities to respond to growing disease burden

- “The human resource should be ready. As you may note, as pathologists, we are already struggling with numbers. We are less than 40 pathologists in the country”
- “In my country we have a short take pathologist as well as a We recruit every year only five residents, but right now up to ten. So we have a shortage. We need to produce more and more pathologists working in the country.”

...█Code: Training healthcare professionals:

Comments related to training healthcare professionals (can be post or within studies) in understanding and using AI.

- “The next point is the training of personnel you know people have been used to individual reading and giving reports now it's going to be most likely taken over by AI and if the people are not well trained it may be bring challenges.”
- “And the training of pathologists we have a unified national master pathology program that trains pathologists and AI is within the curriculum so it it means that they have to adapt to that in in their own training as well as part of the future that they are going to face as pathologists.”

...█Code: Envisioned role of AI:

Vision/ thoughts on how AI would be used in clinical practice, including which use cases (e.g. cancer detection, screening etc), automation level, expected impact (accuracy, speed, access)

- “And I suppose it was stimulated by the fact that there is so, there are so many parameters in assessing a cancer that it was felt that AI could help to put together all the different angles”
- “But if you are going to use it, the AI, to guide the diagnosis or to support the diagnosis, then it's something that is feasible.”

Category: Funding

This category captures perceived sources of funding for AI implementation in cancer diagnostics.

...█Code: Government and public funding:

Mentions regarding funding coming from government/ public institutions (e.g. public hospitals/ health care); Comments on funding/ money/ infrastructure being constrained/ limited, as well as mentions of competition for funding -> related to government

- “the government has to channel funding.”
- “national health insurance system from the government”
- “the ministry of health has to collaborate with finance in order to be able to bring in the money that will finance the infrastructure.”

...█Code: (International) Research funding:

International/ Local research collaborations as sources of funding/ infrastructure provision

- “So there can be collaboration between both the clinical and academic institutions that will also finance that will also prepare the use of this AI assisted modalities in cancer care. And so there can also be, it can even go beyond boundaries between that of us in Ghana, of our colleagues in the Netherlands, or in Uganda or in Malaysia. So this can bring sets of the global-local alliance thereby improving cancer diagnostics and care.”

...█Code: Reimbursement and insurance

Comments regarding insurance systems (public and private), including reimbursement through insurance when patients use clinical services

- “the insurance system can grow to accommodate AI, AI tools, AI diagnostic tools even clinical tool”
- “then there's many, many private insurance. Most people who are employed, salaried, have private insurance through the workplace. But most people also double pay for the government one because it's pretty affordable”

...█Code: Private sector and public-private partnerships

Mentions regarding funding coming from industry/ private healthcare sector (e.g. public-private partnerships, sponsoring equipment, business models)

- “if the companies are ready to provide software, whereby people can do instalment payment it's possible. It's a model which I've been working for many other things including reagent in the labs. Whereby we receive the machine then you buy reagents and continuously become a customer for that.”
- “negotiation with funders so that the cost of hardware is not so prohibitive”

Category: Stakeholder collaborations

...Category: Key stakeholders

Mentions of people/ groups as stakeholders/ relevant groups in the context of pathway steps for integrating AI into cancer diagnostics in LMICs

...█Code: Academic and research institutions

Mentions of university, researchers, scientists, education system, university hospitals, research institutions, academia

...█Code: Clinicians and other health care providers

Mentions of clinicians, oncologists, radiologists, gynaecologists, pathologists, healthcare team, specific people (e.g. head of pathology department) -> overall people, NOT institution/ center/ organizational unit

...█Code: General Public

People, public, students (broad, not specific field)

...█Code: Global health organisations

Mentions of global health initiatives/ organisation

...█Code: Government

Mentions of government, ministries/ ministers, directorates, regulators/ regulatory authority, policy makers, public health insurance

...█Code: Industry / AI vendors

Mention of industry, vendors, companies/ products

...█Code: International research partners

Mention of research groups/ partners in foreign countries

...█Code: Patients

Mention of patients

...█Code: Private healthcare companies

Mentions of private hospital, sector (clearly talking about healthcare), private insurance (companies), private healthcare system

...█Code: Professional medical societies

Mentions of professional bodies, pathology association, scientific association, digital pathology network

...█Code: Public healthcare systems

Hospitals, screening pathway, laboratory, health services, pathology department

...Category: Collaborative structures

...Category: Challenges

......█Code: Fragmentation

Mentions of existence/ danger/ possibility of fragmentation, where people are working on the same project, not planning/ strategizing together (e.g. resulting in interoperability issues)

- “Because we know there's no point of being in isolation. It would just be a lot of repeated effort.”
- “there are different pockets of people”
- “because now it's kind of segmented. There are people who are very good, there are people who are not aware of it and how it can help.”

......█Code: Lack of sustainability

Mentions of danger/ lack of sustainability in collaboration/ partnerships

- “So that's quite a big drawback because sustainability of such can be a problem. And to fully adopt something, we need to make sure that we have spent or provided for its wholesome adoption within the Ministry of Health System”

...Category: Types of collaboration

......█Code: (International) Research collaboration

Mentions of collaboration/ collaborative processes in research (both with local and international partners)

- “the collaboration in the area of innovation, validation of AI tools”

......█Code: Clinical-University

Mentions of collaboration/ collaborative processes between clinics and university

- “So there can be collaboration between both the clinical and academic institutions that will also finance that will also prepare the use of this AI assisted modalities in cancer care.”

......█Code: Global heath organisations-Government/Clinical

Mentions of collaboration/ collaborative processes between global health organisations and local government/ healthcare system

- “And like I mentioned, although it is something that the ministry and university is doing, these are usually supported by partners. They are not local initiatives.”

......█Code: Public-private

Mentions of collaboration/ collaborative processes between industry/ private sector and public sector/ government/ public healthcare

- “private public partnerships”
- “And I think it's possible to vet private systems and make a partnership with a group of them”

......█Code: Advocacy and lobbying

Mentions of lobbying/ bringing in ideas, demonstrating benefit, discussion, advocacy between government and other groups

- “the leadership is closer to the people. So if you want to pitch something in your county, you can easily access the leaders.”
- “Um, even if it's funding government, there are groups that will lobby on the side of pathologists or on the side of doctors on the government side”

......█Code: Key stakeholder meetings

Mentions of meetings between several stakeholders, need for consensus/ identifying mutually beneficial goals/ ways forward

- “And we realized that the next steps, perhaps the next first steps, should be to really get together some kind of consensus as to how we're going to prioritize the focus of developing AI. And I think last year, late last year, we had quite a big meeting together”

......█Code: Interdisciplinary

Mentions of interdisciplinary collaboration (e.g. between different clinicians, clinicians and researchers)

- “Because now once with AI assisted diagnostics there can be better collaboration between the oncologists, radiologists, pathologists.”
